# SARS-CoV-2 epidemic after social and economic reopening in three US states reveals shifts in age structure and clinical characteristics

**DOI:** 10.1101/2020.11.17.20232918

**Authors:** Nathan Wikle, Thu Nguyen-Anh Tran, Bethany Gentilesco, Scott M Leighow, Joseph Albert, Emily R Strong, Karel Břinda, Haider Inam, Fuhan Yang, Sajid Hossain, Philip Chan, William P Hanage, Maria Messick, Justin R Pritchard, Ephraim M Hanks, Maciej F Boni

**Author notes:** equal contribution (Nathan Wikle, Thu Nguyen-Anh Tran).

## Abstract

In the United States, state-level re-openings in spring 2020 presented an opportunity for the resurgence of SARS-CoV-2 transmission. One important question during this time was whether human contact and mixing patterns could increase gradually without increasing viral transmission, the rationale being that new mixing patterns would likely be associated with improved distancing, masking, and hygiene practices. A second key question to follow during this time was whether clinical characteristics of the epidemic would improve after the initial surge of cases. Here, we analyze age-structured case, hospitalization, and death time series from three states – Rhode Island, Massachusetts, and Pennsylvania – that had successful re-openings in May 2020 without summer waves of infection. Using a Bayesian inference framework on eleven daily data streams and flexible daily population contact parameters, we show that population-average mixing rates dropped by >50% during the lockdown period in March/April, and that the correlation between overall population mobility and transmission-capable mobility was broken in May as these states partially re-opened. We estimate the reporting rates (fraction of symptomatic cases reporting to health system) at 96.0% (RI), 72.1% (MA), and 75.5% (PA); in Rhode Island, when accounting for cases caught through general-population screening programs, the reporting rate estimate is 94.5%. We show that elderly individuals were less able to reduce contacts during the lockdown period when compared to younger individuals. Attack rate estimates through August 31 2020 are 6.4% (95% CI: 5.8% – 7.3%) of the total population infected for Rhode Island, 5.7% (95% CI: 5.0% – 6.8%) in Massachusetts, and 3.7% (95% CI: 3.1% – 4.5%) in Pennsylvania, with some validation available through published seroprevalence studies. Infection fatality rates (IFR) estimates for the spring epidemic are higher in our analysis (>2%) than previously reported values, likely resulting from the epidemics in these three states affecting the most vulnerable sub-populations, especially the most vulnerable of the ≥80 age group.

## 1 Introduction

The coronavirus SARS-CoV-2, the cause of coronavirus disease 2019 (COVID-19), has caused significant morbidity and mortality across the United States. During spring 2020, a critical question in managing the United States COVID-19 epidemic was whether regional re-openings of social and economic activity would result in rebounds of cases and hospitalizations [1]. Because population-level immunity to SARS-CoV-2 was still low at the time, the expectation was that increases in mobility and human contact would lead back to an upward trending epidemic curve [2]. However, as hand hygiene, physical distancing, epidemiological awareness, and mask-wearing practices changed during the spring, increases in daily economic and social activity were not guaranteed to recreate the ideal transmission conditions of March. Additionally, no school session and the possible effects of drier/hotter weather [3] in summer were considered potential mitigators on viral transmission [4].

Despite these mitigating factors, epidemiological rebounds had begun in more than 40 states by July 1. Daily case numbers in the US – which had dropped from a peak of 30,000/day in early April to 20,000/day in late May – rebounded to 50,000/day the first week of July [5,6] driven by early re-opening policies in several large states. With a symptomatic case fatality rate (sCFR) rate in the 1% to 4% range [7–11] depending on epidemiological context and testing availability, more than a thousand of these daily new case numbers would result in death several weeks later. The absence of careful, gradual, managed reopenings during the May/June period were the likely cause of summer resurgence in parts of the southern US. It is of utmost public health importance that the next epidemic management milestone – preparation for the 2020/2021 winter wave – is approached with a strategic and adaptive plan that can utilize real-time epidemiological analysis (e.g. attack rate estimates, changing age/mobility patterns, clinical improvements) to contain and potentially reverse upward epidemic trends.

Here, we analyze the age-structured case, hospitalization, and death time series from three states – Rhode Island (RI), Massachusetts (MA), and Pennsylvania (PA) – that during summer 2020 did not experience substantial epidemic rebounds when compared to March/April levels. We evaluate eleven clinical data streams reported by the respective state health departments in a Bayesian inference framework built on an ordinary differential equations (ODE) age-structured epidemic model that includes compartments (clinical states) for hospitalization, critical care, and mechanical ventilation. We infer parameters on surveillance, transmission, and clinical characteristics of the first epidemic wave in RI, MA, and PA. We describe the patterns of persistently low transmission in these three states through August 31, compare these low-transmission scenarios to changes in human mobility metrics, and evaluate changes in age structure and clinical outcomes. We evaluate the impact of the spring epidemic on elderly populations in these three states, and we compare infection fatality rates (IFR) to published estimates from other parts of the world. Preliminary analyses have been posted regularly at https://mol.ax/covid and shared with the respective state departments of health. The statistical inference described here — on attack rates, underreporting, changing age-profiles — can provide improved guidance for real-time decision making and public health messaging.

## 2 Results

### 2.1 Epidemic characteristics during and after lockdown

In Rhode Island, Massachusetts, and Pennsylvania, from early March to early April, we inferred a reduction in the composite parameter *β*_*t*_ describing person-to-person contact (mixing) rates and the probability of virus transmission per unit contact. From the March 5-15 period to April 1, population mixing rates dropped by 56.3% (95% CI: 42.9% - 67.5%) in RI, by 76.6% (95% CI: 66.4% - 85.2%) in MA, and by 89.8% (95% CI: 88.0% - 91.9%) in PA (Figure 1). During this period, contact rates were dropping through stay-at-home orders, bans on large gatherings, and business/school-closures at the same time as improved hygiene behaviors were being increasingly adopted; thus, it is not possible to determine the individual contributions to *β*_*t*_ of mixing reduction and hygiene improvement. The reductions seen in *β*_*t*_ in March and early April are reductions in *transmission-capable mixing* that result both from fewer person-to-person contacts and lower infection risk per contact. Note that in a heterogeneously exposed population, mixing rates for large highly connected groups can drop by large amounts with only a modest drop in the population’s effective reproduction number *R*_*t*_ if a smaller sub-population maintains a chain of infection due to an inability to completely zero-out contacts. For example, if 90,000 office employees can work from home and contact only their families but 10,000 elderly care home residents still require contact with medical and care staff, then a full business shutdown may result in a 90% reduction in mixing patterns but an *R*_*t*_ ≈ 1 if a stable chain of infection is maintained in nursing homes and elderly care residences. Our estimated reductions in transmission-capable mixing are consistent with published estimates of changes in *R*_*t*_ and mobility [12–16].

**Figure 1.**
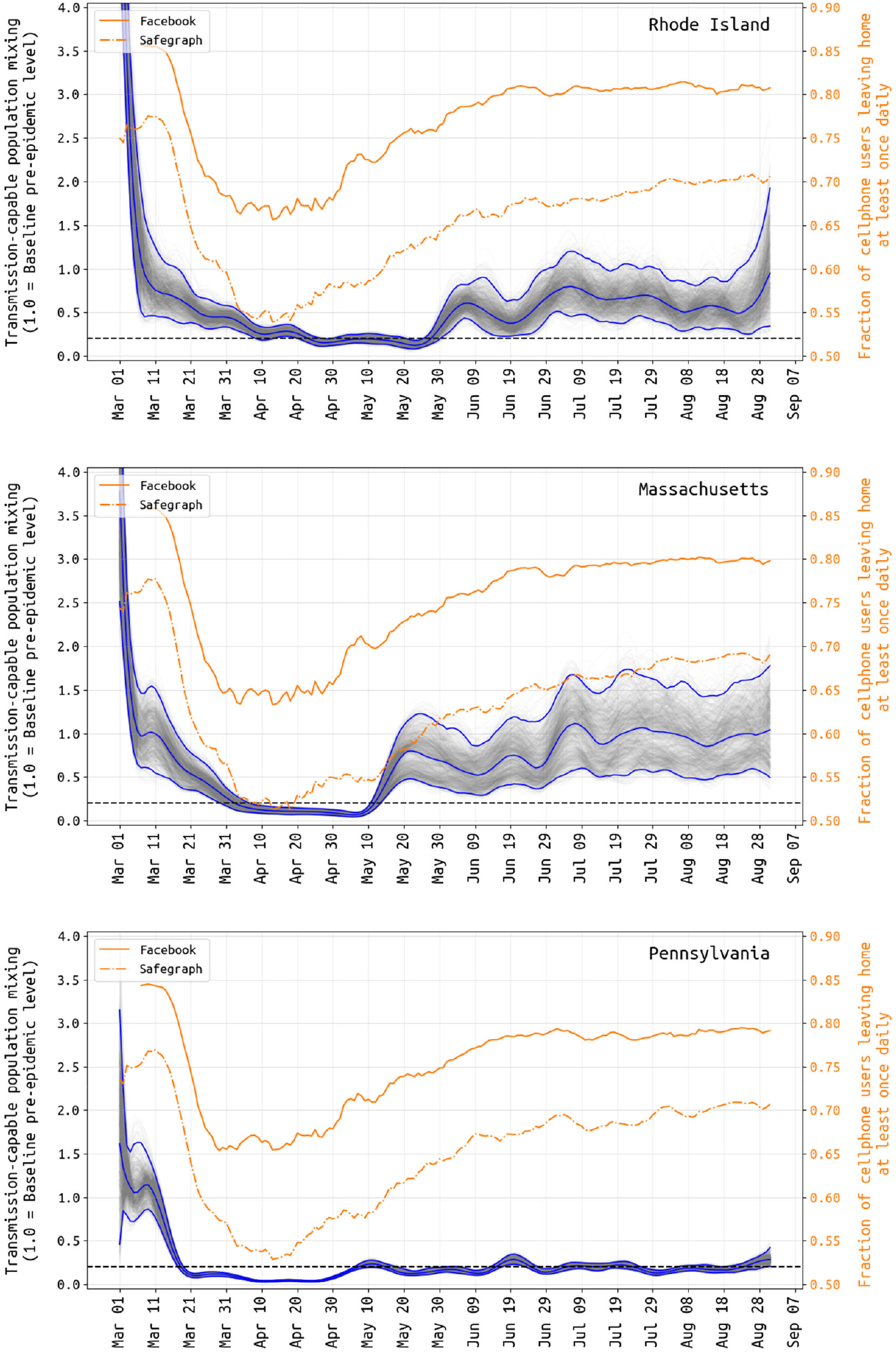
Transmission-capable mixing *βt* (in gray and blue) and mobility changes (yellow) from March 1 to August 31. The average population mixing for March 5-15 is set to 1.0 as the pre-epidemic level of transmission-capable mixing, and all other values are reported relative to this. Gray lines show 1000 sampled posterior *β*-trajectories with the blue lines showing the median and 95% credible intervals. Note that there is substantial uncertainty in these estimates during the first weeks of March, as case numbers were low and reporting may not have been catching a large proportion of true cases at this time. Yellow lines show the fraction of Facebook and SafeGraph users that left home at least once per day. The correlation between population-movement (yellow) and transmission-capable population movement (gray+blue) begins to disappear in early May in all three states.

**Figure 2:**
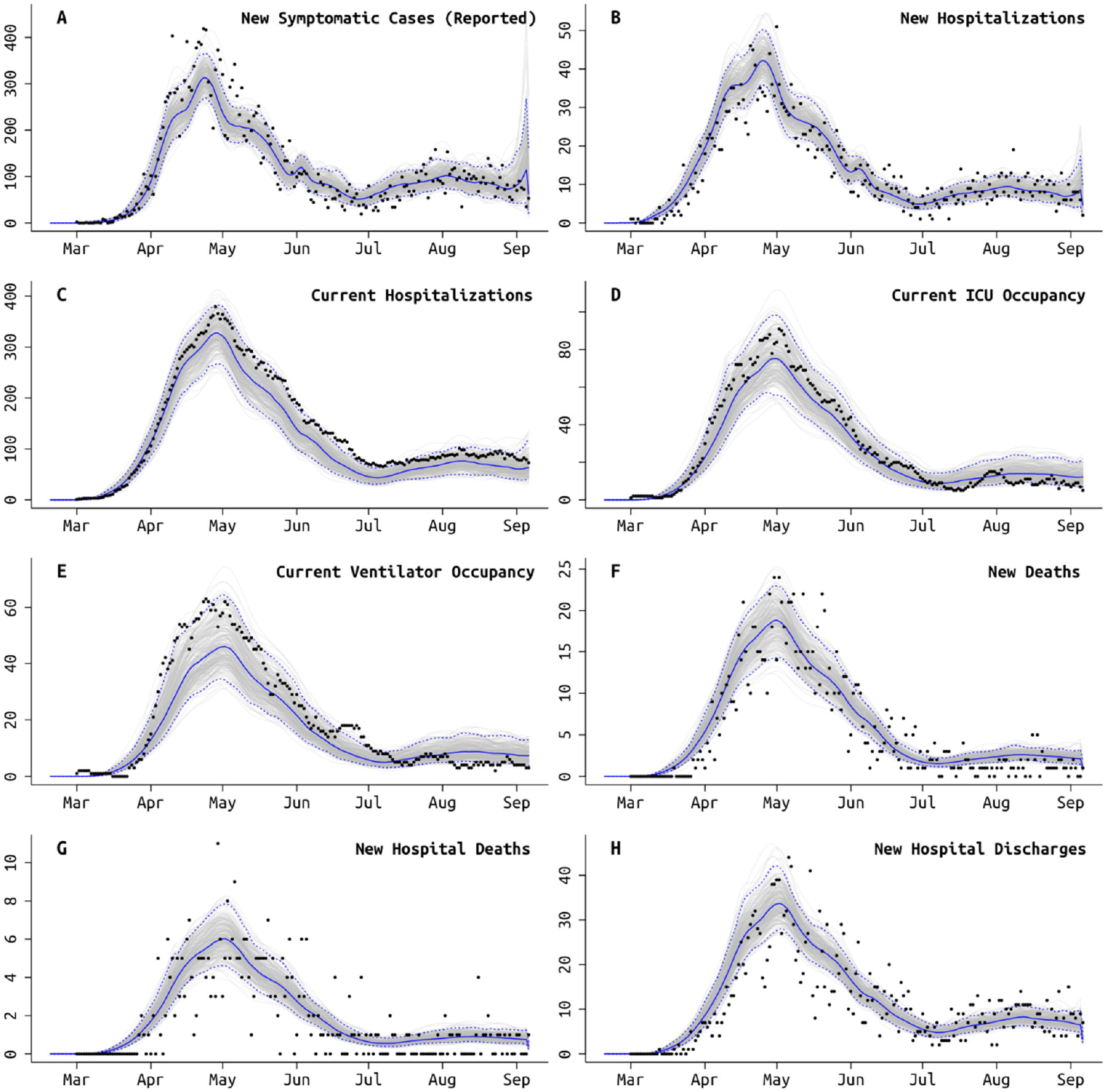
Model fit to Rhode Island daily data, using the best fit model which accounts for different age-based contact rates after the lockdown and a different rate of ICU admissions starting in early June (Table 3). Gray lines show 250 sampled trajectories from the posterior, and blue lines are the median trajectories. Black circles are data points that show the daily (**A**) newly reported symptomatic cases, (**B**) new hospitalizations, (**C**) current number of patients hospitalized, (**D**) current number of patients in critical care, (**E**) current number of patients undergoing mechanical ventilation, (**F**) new deaths reported, (**G**) new hospital deaths reported, i.e. excluding deaths that occurred at home or at long-term care facilities, and (**H**) number of hospital discharges. Model fits for MA and PA are shown in Figures S3 and S4.

Changes in the inferred population-mixing component *β*_*t*_ can be compared to mobility metrics [17,18] based on location-enabled smartphone data trails, which allow calculation of time spent at home versus outside the home. Two independent mobility data sources, Facebook and SafeGraph [19,20], provided daily estimates for the fraction of tracked users leaving home at least once in a 24-hour period. Despite values varying slightly across states and significantly between user bases, all mobility data examined have a common shape and timing: an initial baseline in early March (84%-86% of users leaving home for Facebook; 75%-77% for SafeGraph), and a subsequent dramatic decrease from 3/15 to 3/31 (64%-67%; 52%-55%). This low fraction of users leaving home (at a minimum, once daily) is maintained until about April 20, followed by a slow increase to a slightly more cautious ‘new normal’ (77%-81%; 66%-69%) through July and August (Figure 1). A resumption of population mobility in early May suggests that improvements in hygiene, personal distancing, mask wearing, selective travel, and/or smaller event sizes were likely factors keeping *R*_*t*_ < 1 and new case numbers declining.

Not all symptomatic SARS-CoV-2 infections are reported to state-level health systems. As it is difficult to make distinctions among asymptomatic, sub-clinical, mildly symptomatic, and symptomatic infections, here we call an infection *symptomatic* if the symptoms are pronounced enough that a person with convenient zero-cost access to health care would choose to visit a hospital or clinic. Using the delays between time series of cases, hospitalizations, and deaths, we can estimate the fraction *ρ* of symptomatic cases that are reported to the health system [21,22]. We do this without making assumptions about the case fatality rate or infection fatality rate. However, two complications present themselves at state-level reporting systems in the US. First, new hospitalizations (incidence) can also be underreported (as in PA), and second these numbers may be available only as current hospitalizations rather than new or cumulative hospitalizations (as in MA). When hospitalization incidence is underreported as in PA, the case-and-hospitalization time series needs to be statistically linked to a data stream that is complete in its coverage of the general population (normally deaths). In Pennsylvania we estimate that 77.0% (95% CI: 62.8% - 87.1%) of hospitalized cases were reported daily on the PA DOH website, and our estimate of the symptomatic case reporting parameter *ρ* is 75.5% (95% CI: 61.7% - 91.8%). When only current hospitalizations are available (MA), a good model fit requires that the duration of hospitalization is known or identifiable; this is complicated by the fact that hospital stays come in several categories (admission to ICU, requiring mechanical ventilation) and can be censored by death events. In Massachusetts, there is not enough information in the remaining data streams to identify the duration of hospitalization, and as a result an externally validated assumption is made (prior distribution 11.8–12.8 days, based on [23]); age-stratified probability of hospital admission in MA is constrained to be close to estimated values obtained from Rhode Island data (Figure 5E). Our estimate for the reporting rate *ρ* in Massachusetts is 72.1% (95% CI: 60.4% - 84.5%). Rhode Island has complete reporting of hospitalization incidence, made possible by the state’s small size and a reporting system covering several small hospital networks that include all hospitals in the state. We estimate that 96.0% (95% HPD: 84.2% - 99.8%) of symptomatic COVID-19 cases are reported to RIDOH (after May 2020). RIDOH staff and affiliated physicians reported that patients were being turned away in early March due to lack of tests, and March reporting rates are estimated at less than 30% (March 15 estimate is 13.2%, 95% CI: 7.4% - 26.1%); see Figure 5A.

A second complication in estimating a state-level surveillance system’s reporting rate is that the daily presented case numbers combine cases from both passive surveillance and random screening of asymptomatic individuals. The reporting rate *ρ* should be adjusted downward when levels of asymptomatic random screening are known and high, a correction that is not included in current estimation methods [22]. In Rhode Island, monthly screening numbers are available (Supplementary Materials, Section 3.7) and indicate that 40% of diagnostic tests administered were done so in congregate care settings, in business settings for high-contact individuals, known contacts of cases, and other settings where it was deemed helpful to test asymptomatic individuals. Despite this being a large proportion, only a small fraction (typically <1%) of randomly screened individuals test positive in these settings. Adding the monthly random screens into our likelihood framework moves our estimate of Rhode Island’s reporting rate down to 94.5% (77.5% – 99.8%). With higher levels of asymptomatic random screening, a recommended cornerstone for continued COVID-19 epidemic management [24], the bias in inferring *ρ* may be substantial. Numbers of monthly random screens are not available in PA or MA.

Reporting rate estimates combined with age-specific estimates of asymptomatic infection [25] allow cumulative attack rates to be estimated (Figure 3). The probability of asymptomatic infection for SARS-CoV-2 requires prospective follow-up in either a household or cohort design, with few studies including enough age groups for between-age comparisons [26–29]. We compared (*a*) a basic weighted-average across studies with sufficient follow-up to distinguish pre-symptomatic from asymptomatic (see Supplementary Materials Section 1.8), (*b*) recently published estimates from Davies et al [25] using model-fitting to both symptomatic time series and cohort data with identified asymptomatics, and (*c*) a set of age-invariant asymptomatic fractions ranging from 0.3 to 0.7. These seven hypotheses were evaluated in our Bayesian model (Table 2) with equal prior weight. The majority of posterior support pointed to “60% symptomatic”, with substantial support for the age-proportions in Davies et al [25]; the Davies proportions were used in all final runs as they were estimated externally. The August 31 population attack rates for SARS-CoV-2 are 6.4% in Rhode Island, 5.7% in Massachusetts, and 3.7% in Pennsylvania (95% CI shown in Figure 3). These attack rate estimates use symptomatic case data through September 6, as an infection on August 31 would have its mean time of symptoms occurrence six days later. The Rhode Island attack rate is able to be validated with a 2.2% late-April attack-rate estimate obtained from a household sero-survey [30] and 0.6% early-April estimate from blood donors [31] (population biased towards healthier individuals). Our Pennsylvania-wide attack rate has a Philadelphia early-April estimate of 3.2% as a comparator [32], as well as a 6.4% estimate from July using serum from dialysis patients (not adjusted for race or socio-economic indicators, and thus biased upward) [33]. The unadjusted dialysis-patient seroprevalence in Massachusetts was estimated 11.3% in July 2020 [33], about twice our estimate.

**Table 1.** Infection fatality rate (IFR), symptomatic case fatality rate (sCFR), and hospitalization fatality rate (HFR) for the March-May COVID-19 epidemics in RI, MA, and PA. Numbers of deaths observed in the <20 age groups were too low to generate meaningful estimates. CI: credible interval.

| Age Range | State | IFR (95% CI) | sCFR (95% CI) | HFR (95% CI) |
| --- | --- | --- | --- | --- |
| 20-29 | RI | <5 deaths | <5 deaths | <5 deaths |
|  | MA | 0.03% (0.03% - 0.04%) | 0.13% (0.10% - 0.16%) | 4.0% (3.4% - 5.0%) |
|  | PA | 0.03% (0.03% - 0.04%) | 0.13% (0.10% - 0.16%) | 4.9% (4.3% - 6.0%) |
| 30-39 | RI | 0.06% (0.04% - 0.07%) | 0.17% (0.13% - 0.21%) | 2.8% (2.2% - 3.9%) |
|  | MA | 0.08% (0.07% - 0.10%) | 0.24% (0.20% - 0.29%) | 4.3% (3.6% - 5.3%) |
|  | PA | 0.07% (0.06% - 0.09%) | 0.21% (0.19% - 0.27%) | 5.3% (4.6% - 6.4%) |
| 40-49 | RI | 0.30% (0.22% - 0.42%) | 0.76% (0.56% - 1.06%) | 9.1% (6.5% - 12.2%) |
|  | MA | 0.47% (0.39% - 0.53%) | 1.18% (0.97% - 1.32%) | 14.6% (12.0% - 16.3%) |
|  | PA | 0.49% (0.42% - 0.61%) | 1.23% (1.04% - 1.52%) | 19.3% (16.2% - 22.4%) |
| 50-59 | RI | 0.58% (0.40% - 0.77%) | 1.18% (0.81% - 1.58%) | 9.6% (6.8% - 12.8%) |
|  | MA | 0.80% (0.67% - 0.90%) | 1.64% (1.37% - 1.85%) | 15.4% (12.7% - 17.1%) |
|  | PA | 0.94% (0.80% - 1.17%) | 1.92% (1.61% - 2.36%) | 20.4% (17.1% - 23.6%) |
| 60-69 | RI | 3.2% (2.5% - 4.2%) | 5.2% (3.9% - 6.6%) | 13.3% (9.4% - 17.7%) |
|  | MA | 2.7% (2.2% - 3.0%) | 4.3% (3.5% - 4.8%) | 21.2% (17.4% - 23.6%) |
|  | PA | 2.9% (2.4% - 3.6%) | 4.6% (3.8% - 5.7%) | 28.1% (23.6% - 32.5%) |
| 70-79 | RI | 12.0% (9.4% - 14.0%) | 17.3% (13.6% - 20.3%) | 23.2% (18.7% - 30.7%) |
|  | MA | 11.9% (9.8% - 13.7%) | 17.3% (14.1% - 19.9%) | 33.6% (28.7% - 41.0%) |
|  | PA | 8.8% (7.1% - 10.8%) | 12.8% (10.3% - 15.6%) | 40.6% (35.5% - 49.2%) |
| ≥80 | RI | 20.1% (15.8% - 24.0%) | 29.2% (22.9% - 34.8%) | 27.3% (22.6% - 35.5%) |
|  | MA | 22.9% (18.9% - 25.3%) | 33.1% (27.4% - 36.7%) | 38.7% (33.4% - 46.6%) |
|  | PA | 17.3% (14.0% - 20.6%) | 25.0% (20.3% - 29.8%) | 46.2% (40.7% - 55.5%) |

**Table 2.** Posterior probabilities of seven different assumptions on asymptomatic infection. After a million iterations, samples of 1000 from posterior.

|  | 30% symptomatic all-ages | 40% symptomatic all-ages | 50% symptomatic all-ages | 60% symptomatic all-ages | 70% symptomatic all-ages | Simple average in 20-yr age bands, excluding studies with “pre-symptomatic infections” | Davies et al asymptomatic proportions |
| --- | --- | --- | --- | --- | --- | --- | --- |
| RI | 0.008 | 0.070 | 0.207 | 0.474 | 0.238 | 0.003 | 0.0 |
| MA | 0.0 | 0.004 | 0.023 | 0.106 | 0.112 | 0.127 | 0.628 |
| PA | 0.0 | 0.001 | 0.0 | 0.0 | 0.0 | 0.0 | 0.999 |

**Figure 3.**
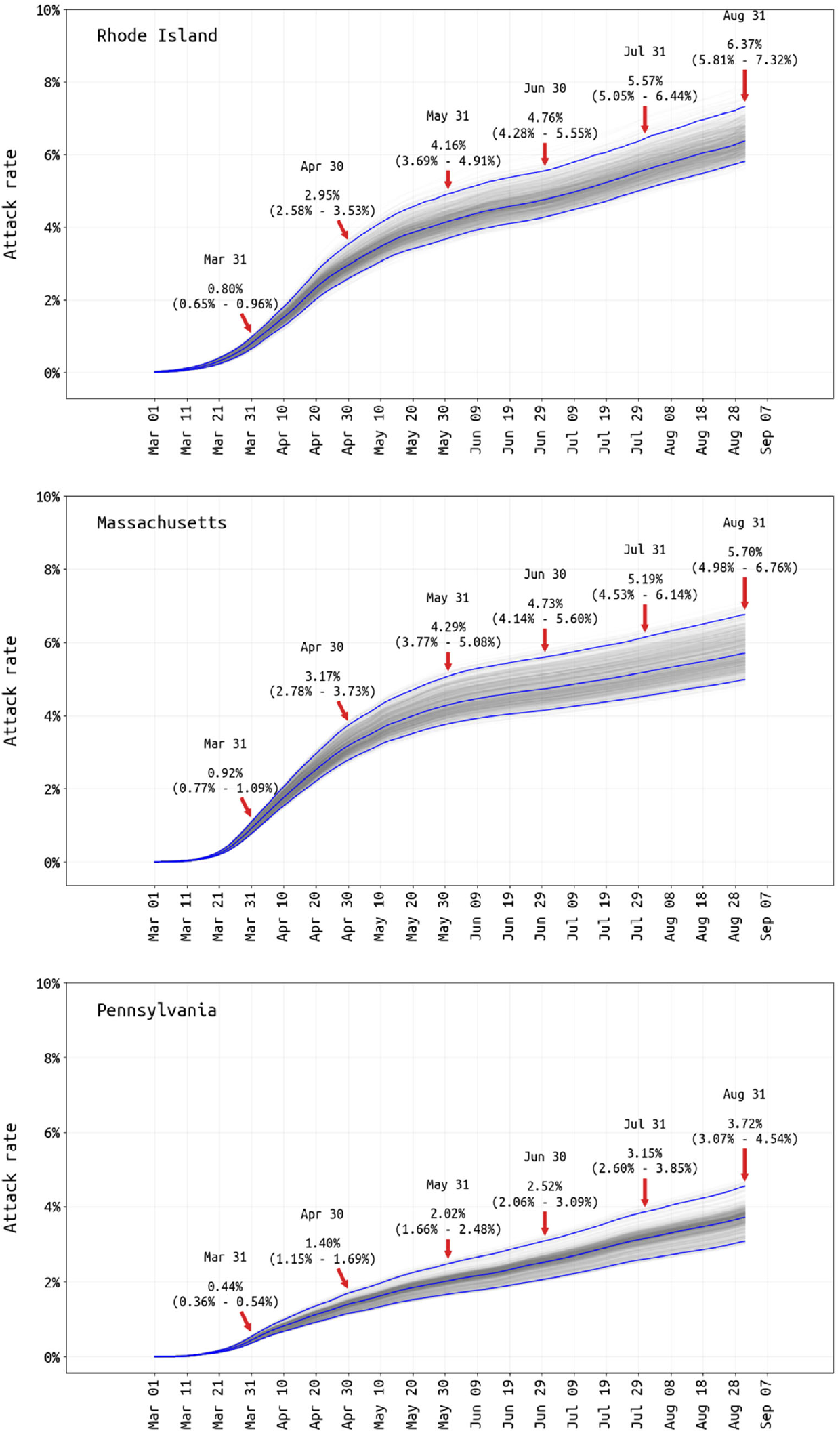
Posterior distribution of total attack rate through August 31 2020. Total infection attack rate includes all reported symptomatic cases, estimated unreported symptomatic cases, and estimated asymptomatic cases. Cumulate attack-rate estimates and 95% credible intervals are shown for the end of every month.

With marked increases in cases occurring in all three states in October 2020, the October 31 attack rate estimates (using data through Nov 6) are 10.9% (95% CI: 9.8% - 12.6%) in Rhode Island, 8.2% (95% CI: 7.8% - 8.7%) in Massachusetts, and 6.8% (95% CI: 6.4% - 7.4%) in Pennsylvania.

Estimates of attack rate allow for age-specific infection fatality rate (IFR) estimation (Table 1). First, our results show that the age-adjusted March-to-May IFR for all three states is higher than the typically quoted 0.5%–1.0% range over the past eight months of IFR-estimation [34], but note that epidemics that infect the most vulnerable segments of a population first may be associated with higher-than-average IFRs (see *Discussion*). IFR estimates are 2.5% (95% CI: 1.9% – 2.8%) for the early Rhode Island epidemic, 2.3% (95% CI: 1.9% – 2.6%) for the early Massachusetts epidemic, and 2.1% (1.7% – 2.5%) for the early Pennsylvania epidemic. Estimated IFRs for the entire epidemic are somewhat lower (especially in PA) due to a lower rate of ICU admissions in summer (see below). It is well known that the IFR depends strongly on age, gender, co-morbidities, socio-economic factors, and race [35,36]. Our estimated age-stratified IFRs indicate that fatality rates are highest (>2%) in the 60+ age groups, still very high in the 40-59 age group (estimates ranging from 0.30% to 0.94%), and lower in the <40 age group (<0.1%). The age-adjusted symptomatic case fatality rates (sCFR) are estimated to be 3.7% (RI), 3.5% (MA), and 3.2% (PA), all higher than the national sCFR for the United States. The hospitalization fatality rate (HFR) shows the least variation by age, with fatality rates >9% for the >40 age groups, 2.8% to 5.3% HFR for 20-39 age group, and no estimates possible for individuals under 20.

### 2.2 Changes in age-stratified contact patterns and clinical outcomes during the epidemic

We investigated changing age-specific contact rates during the three state epidemics, based on observed changes in age distribution and well-documented reporting of outbreaks in nursing homes. Since age-contact matrices have only recently begun to be measured for the COVID-19 socially-distanced era [37–41], we infer eight relative mixing levels for each age class (relative to the 0-9 age group) and assume that age-matrix contact rates are the products of individual age-group mixing rates (see *Discussion*). Age-specific mixing patterns are allowed to change when the lockdown ends. Age characteristics of each state epidemic are shown in Figure 4 (*top row*), and the inferred contact parameters are shown in Figure 4 (*middle and bottom rows*); inference of contact rates is influenced by the model assumption that the 0-19 age group is 60% as susceptible to infection as the other age groups [25]. In all three states, the lowest inferred contact rates during lockdown were for the 0-9 and 60-79 age groups, reflecting closed schools and possibly the caution with which older individuals approached their risk of infection. However, the relative contact rates for individuals in the ≥80 age group were much higher: 2.5 (95% CI: 2.3–2.8) in RI, 5.6 (95% CI: 4.8–6.9) in MA, and 8.5 (95% CI: 7.9–9.6) in PA. This suggests that social distancing and lockdown were more difficult for individuals that needed additional care or lived in congregate care facilities. The shift from an older age profile to a younger age profile is apparent in all three states’ epidemics as the epidemics progressed from spring to summer (Figure 4, *top row*). Changes in age and clinical patterns (see below) are supported by lower DIC values (Table 3).

**Table 3.** Deviance Information Criterion (DIC) values for different models. Minimum DIC values shown in boldface.

|  | No change in age-profile of contact rates post lockdown. | Different age-profile of population contact rates post lockdown (approx May 2020). | No change in age-profile of contact rates post lockdown. | Different age-profile of population contact rates post lockdown (approx May 2020). |
| --- | --- | --- | --- | --- |
|  | No change in ICU admission rate from March to August. | No change in ICU admission rate from March to August | Allows for change in ICU admission rate from March to August. | Allows for change in ICU admission rate from March to August. |
| RI | 14427 | 13443 | 14835 | **13407** |
| MA | 29773 | 23436 | 29297 | **22964** |
| PA | 25103 | 14924 | 24837 | **14793** |

**Figure 4.**
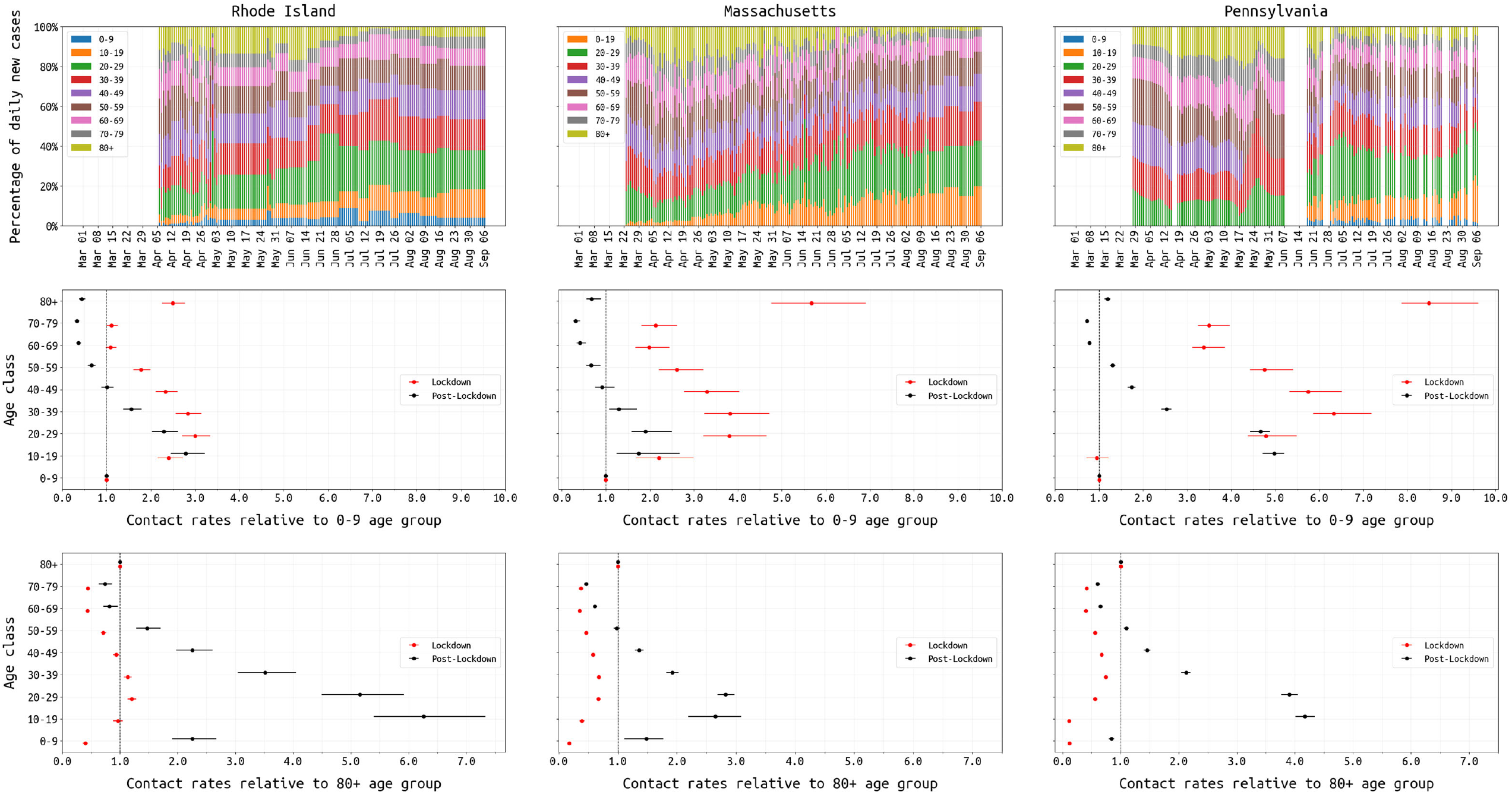
Changing age-structure of COVID-19 epidemics in RI, MA, and PA. Top row shows the age structure of reported cases from March 1 to August 31. RI and PA report age data periodically; missing values have been linearly interpolated in RI. Middle row shows the inferred age-specific contact rates (median and 95% credible intervals) for both the lockdown (red) and post-lockdown period (black), where the reference group is the 0-9 age group. Bottom row shows the same inferred contact rates as the middle row with the ≥80 age group as the reference.

**Figure 5.**
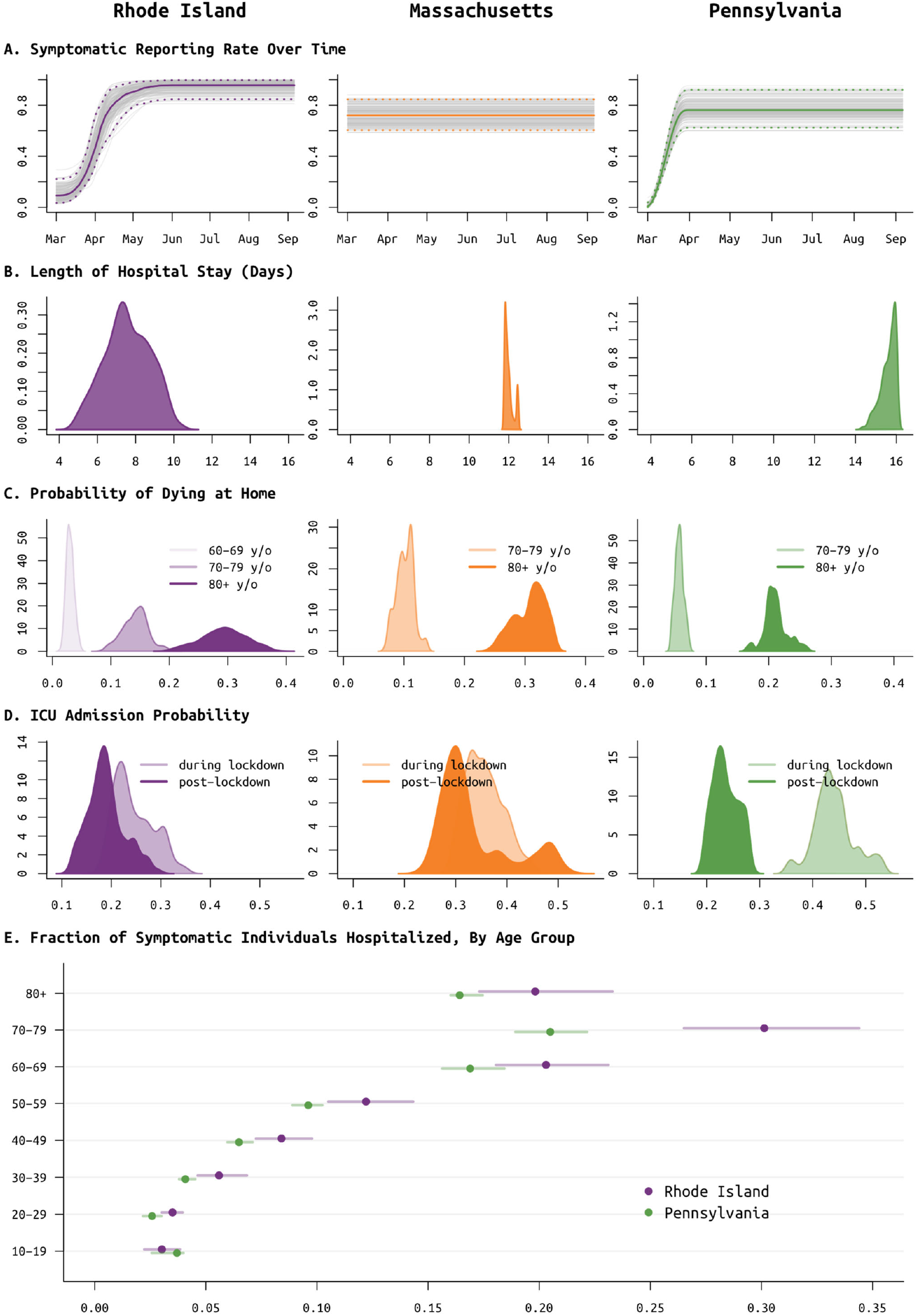
Posterior distributions of reporting rate (panel **A**) and clinical parameters (panels **B** to **E**) for Rhode Island (purple, left column), Massachusetts (orange, middle column), and Pennsylvania (green, right column). (**A**) Reporting parameter *ρ*, i.e. the fraction of symptomatic SARS-CoV-2 cases that are reported to the health system, plotted as a function of time. In Rhode Island, it was known that in March testing was not available and cases could not be confirmed; therefore a spline function was fit for *ρ*. This same function provided a better fit for Pennsylvania data, but not for Massachusetts data. (**B**) Median length of medical-floor hospital stay was 7.5 days in RI, 11.9 days in MA, and 15.7 days in PA. This parameter was constrained to be between 11.8 and 12.8 days in MA, as without this constraint identifiability issues arose due to the lack of the ‘cumulative hospitalizations’ data stream. (**C**) Probabilities of dying at home for the 60-69, 70-79, and 80+ age groups; 60-69 age group was included only for RI as data were insufficient in PA and MA. These are largely reflective of the epidemics passing through nursing home populations where individuals are not counted as hospitalized if they remain in care at their congregate care facility in a severe or advanced clinical state. These probabilities are important when accounting for hospital bed capacity in forecasts. (**D**) Age-adjusted ICU admission probability during the lockdown period in spring 2020 (lighter color) and after the lockdown (darker color). (**E**) Probability of hospitalization (median and 95% CIs) for symptomatic SARS-CoV-2 infections, by age group; MA estimates are excluded as these had priors set based on estimates in RI.

Improvements in clinical management of hospitalized COVID-19 cases, due to the use of prone positioning [42,43] or more frequent use of corticosteroids [44,45], may have led to lower mortality relative to epidemic size during the more recent (June-Aug) stages of the epidemic when compared to March/April mortality rates [46–48]. To estimate the effects of some of these interventions, we assess whether progression from hospitalization to critical care changed between the early stages and the later stages of the epidemic. Our model uses the relative age proportions described by Lewnard et al [23] who estimated probabilities of progression from medical-floor care to critical care to be between from 30% to 50% (see also [49,50]) for all nine age bands used in this study. These age-specific probabilities are scaled in our model keeping the relative age probabilities the same; scaling is independent for each state as patterns of hospital admission and clinical algorithms for ICU admission are likely to differ somewhat between health systems and hospitals. In Rhode Island, the age-adjusted probability (posterior median) of ICU admission for a hospitalized case dropped from 23.5% (95% CI: 18.8%–32.9%) to 18.5% (95% CI: 12.6%–27.3%) with an inferred breakpoint at June 2 2020 (95% CI: April 11 – July 6). In Pennsylvania, the age-adjusted ICU admission probability dropped by a factor of two from 43.5% (95% CI: 36.1% – 52.6%) to 23.2% (95% CI: 19.6%– 27.9%), with an inferred breakpoint of June 21 (95% CI: June 3 – June 29). In Massachusetts, this probability dropped from 35.3% (95% CI: 29.7% – 43.6%) to 30.9% (95% CI: 25.5% – 49.4%), with the likely change occurring between late-May and mid-June (Figure 5D). In Massachusetts, the lack of a ‘cumulative hospitalizations’ data stream leads to less reliable MCMC convergence, giving us less confidence in the statistical signal showing a reduction in ICU admissions through time.

A second approach to confirming trends on improved clinical case management would be to look directly at changes in mortality. However, the complexity in this analysis lies in the different possible clinical paths that lead to a fatal outcome. In most states, reported mortality trends combine deaths occurring in hospitals with deaths occurring at home (i.e. in congregate care facilities); these data streams are separated in RI/PA but not MA. Our model allows for inference of at-home mortality, with the at-home symptomatic case fatality ratio estimated at between 20% to 35% for the ≥80 group, and 5% to 15% for the 70-79 age group (Figure 5C). This allows us to separate mortality trends into home and hospital, but hospital mortality alone is a complex composite of probability of death on the medical-floor level of care and probability of death in the ICU (with and without ventilation). For this reason, we chose ICU admission as the clinical progression marker where we could evaluate a simple trend of improved case management.

## 3 Discussion

This is among the first studies to evaluate a large set of multiple simultaneous clinical data streams with an epidemic transmission model. The analysis of concurrent data streams is necessary to describe certain important but unreported characteristics of regional SARS-CoV-2 epidemics; these include underreporting of cases, underreporting of hospitalizations, changing age patterns of infection, changing patterns of clinical progression, an understanding of mortality rates outside hospital settings, and investigation of population heterogeneities in susceptibility, vulnerability, and mortality. The inclusion of multiple age-structured data streams on death and hospitalization allows for statistical estimation of symptomatic case underreporting — a quantity that is generally resistant to robust estimation especially in public-health reporting systems that (1) mix active and passive surveillance, (2) mix multiple diagnostic tests and testing visits, and (3) have not made estimates of their catchment areas. With an estimate of symptomatic case underreporting (here, via *ρ*), we can estimate the population-level SARS-CoV-2 attack rate in each state by summing the reported symptomatics, the unreported symptomatics, and an externally estimated number of asymptomatics. One month later, an attack rate estimate can be validated by comparing to results from a seroprevalence survey. Four seroprevalence estimates available for RI, MA, and PA show no major inconsistencies with our results. It is important to remember that SARS-CoV-2 sero-surveys can be subject to biases depending on approaches to recruitment (which can overestimate seropositivity if enriched for individuals who are more likely to have been infected, e.g. individuals who consent because of past symptoms), the time since the original infection (antibody titers have been found to wane over time), and the specific test used [51].

Our results indicate that Rhode Island, Massachusetts, and Pennsylvania are nearly fully susceptible to a winter epidemic wave of SARS-CoV-2. Continual attack-rate estimation will be crucial for the winter’s epidemic response and any planned roll-out of additional public health strategies in spring and summer 2021. Specifically, real-time age-specific attack rate estimation will be important for vaccine planning when vaccination rolls out in early 2021, as age groups experiencing the least infection may need to be accounted for during prioritization of vaccination rollout.

Parallel to attack-rate estimation, mobility tracking can give us a partial window into the effect that distancing policies or lockdowns are likely to have on viral transmission. In May 2020, the positive correlation between stay-at-home metrics and viral transmission vanished in all three states (as in [52]), resulting in a summer with population mixing levels (i.e. people not staying at home) at nearly pre-pandemic levels but viral transmission still at its post-lockdown low point. It is reasonable to suggest that at least some of this is explained by (1) weather increasing the proportion of contacts made outdoors, where transmission is known to be much less likely, and (2) a shift from mixing outside the home to inside the home, i.e. less time spent at work and more family gatherings. It is not straightforward to relate measures of population movement to opportunities for transmission, for many reasons including the collinearity of mixing with many other factors that can influence it. Essentially, rather than absolute measures of mixing, the blue lines in Figure 1 can be interpreted as levels of population mixing that are capable of producing transmission (“transmission-capable mixing”). By cancelling large events, promoting stringent hygiene measures, requiring masking, closing schools, restricting gathering sizes, and creating new guidelines for business operations, the epidemics in RI, MA, and PA were contained during the summer months while allowing the states’ residents to continue most essential activities including small/medium outdoor events. In summer 2020, aggregate measures of population movement were at or near normal levels but mixing leading to transmission was substantially reduced. If lockdown-like measures are required during winter 2020-2021, mobility metrics will allow us to compare stringency between the winter lockdowns and the March/April lockdowns, but macro mobility trends will not be able to capture increased mixing that occurs inside the home.

In our analysis, infection fatality rates are estimated to be higher than in a recently summarized meta-analysis [34], and the differences are particularly notable in the 60-79 age group where we infer IFRs that are 1.5 to 2.5 times as high for the March-to-May spring epidemic waves. Our IFR estimates, ranging from 2.1% to 2.5%, are not however out of range of some of the highest estimates [9] of the Levin et al study [34] and are likely to correspond to a high degree of exposure heterogeneity in the studied epidemics. As it is known that RI and MA had substantial outbreaks in elderly care facilities in the spring, it is likely that this focused epidemic passed through a more susceptible sub-population (individuals who cannot fully quarantine or distance due to needing routine care) that is also more likely to progress to severe clinical outcomes including death. As in all studies to date, our estimates are of ‘epidemic IFR’ and not ‘population IFR’, that is, they are estimates of infection fatality rates actually experienced during an epidemic (which may have run through the most vulnerable populations) and not the fatality rates that would be experienced if the epidemic had run through the entire population or had an equal chance of infecting everyone. As the epidemics continue through subsequent waves of infection, the 2021 and 2022 IFRs will need to be re-estimated and they may prove to be lower. Pennsylvania’s summer 2020 IFR (∼ 1%) was already estimated to be lower than the PA spring IFR (∼2%).

The symptomatic case fatality ratios (sCFR) inferred for RI, PA, and MA (estimates range from 2.5% and 3.7%) are in the higher ranges of previously reported estimates [8–11,53–55], suggesting that the individuals infected during the spring wave and summer lull were more likely to progress to symptoms than the average person in the population. Again, this is consistent with the observation that children were the least exposed in the spring and summer months, and thus the exposed population was both more likely to progress to reportable symptoms and more likely to progress to severe clinical outcomes.

As recent policy discussions (Oct 2020) have been diverted by the capitulation and laziness of an epidemic management approach that would encourage younger/healthier populations to become infected [56], we should restate that our state-level analyses indicate that older individuals are not able to fully isolate during lockdown periods. This makes a ‘protecting the vulnerable’ strategy unworkable, as more vulnerable individuals will still require essential care and contact with other humans. Any policy aiming to protect vulnerable individuals while allowing the remainder of the population to mix and move freely would almost certainly fail at preventing viral introduction from the general population into vulnerable populations. In our analysis, during the March/April lockdown period, the ≥80 contact rate was the highest or among the highest when comparing across age groups (Figure 4). As individuals in the oldest age groups are relatively unaffected by lockdown, the best way to protect these (and other) vulnerable populations is to limit the spread in the general population.

### 3.1 Limitations and Recommendations

One key limitation in using data streams rooted in symptomatic case reporting is the inability to infer asymptomatic infection rates. These rates must be estimated independently from cohort follow-up or contact tracing. They are susceptible to bias in the younger age groups if children test negative due to low viral loads and are classified as negative rather than asymptomatic. Studies are also susceptible to design errors when the protocol or data collection does not allow for differentiation between pre-symptomatic and asymptomatic individuals (Supplementary Materials, Section 1.8). Although the majority of studies have converged on an age-adjusted “60% symptomatic” number, age-specific estimates come with less certainty and differences in diagnostic tests and testing protocols have resulted in substantial variation in these estimates (Supplementary Materials, Figure S1).

The data streams we present here do not allow us to evaluate the degree to which the epidemic runs through specific sub-populations (e.g. congregate care settings, college students) that are more vulnerable, susceptible, or transmit more easily. To measure variability in transmission and susceptibility from state-level data, we suggest including these common data types into the same databases/datasheets currently maintained by all state DOHs as part of routine COVID reporting: (1) contact counts and positivity rates from contact tracing efforts, (2) positive/negative case counts and inclusion criteria from asymptomatic random screening programs, and (3) a datasheet keyed on a categorical variable of ‘infection source event’ with confirmed patient counts, ages, and dates of reporting listed [57,58]. Among these data types, the asymptomatic screening efforts are likely the easiest to turn into a standardized daily data stream as samples taken from screening programs pass through the same sample/data processing pipelines as samples from symptomatic patients. These data would also allow for real-time tracking of prevalence. Random screening efforts in Rhode Island may already be altering our estimation of the reporting rate *ρ*, and the magnitude of this effect is likely to increase in 2021.

We cannot exclude the possibility that our reporting rate estimate (*ρ*) is wrong due to model misspecification or data integrity problems. This is the reason that validation with seroprevalence estimates is crucial for estimating underreporting in public health surveillance systems. The entire inferential framework for *ρ* assumes that hospitalization data are complete (or that hospital underreporting is identifiable), that death data are complete, and that various measures of hospitalization duration have been independently estimated or are identifiable from our data. The biggest leap in these assumptions comes in the completeness of hospitalization data, as both Massachusetts and Pennsylvania have relied on hospitalization data streams that are partially complete. Although the underreporting was able to be estimated in PA, in MA this is not possible because the only cumulative hospitalization numbers are from a followed sub-group of patients that also reported with symptoms. This is a reminder — during the pursuit of rapid results with pre-packaged epidemiological tools and dashboards — to carry out the somewhat slower due diligence of understanding the sampling frames of all data streams included in an analysis. Hospitalization numbers may be underreported in other states as well, and this would mean that national-level analyses of hospitalization numbers would need to acknowledge and account for this. The reporting rate *ρ* determines the attack-rate estimates; if *ρ* is overestimated the infection fatality rates presented here would also be overestimated.

Finally, we were not able to use any published contact matrices for the lockdown period as these data did not exist at the time our work was being done. Thus, we used nine independent mixing rates for the nine age-classes in our model (and assumed that contact between two age groups is proportional to their two mixing rates); the data are unlikely to have enough resolution to infer 81 independent mixing parameters. As confidence grows in the survey methods used to collect post-pandemic age-contact data [37–41] these matrices should be included into modeling efforts like ours.

Infectious disease epidemiology has been neglected in the United States for more than half a century because of our status as a developed country, with a secure food supply, a sanitary water system, few persistent disease vectors, high public hygiene standards, and ample supply of therapeutics and vaccines. We were not prepared in 1981 when the HIV epidemic was uncovered, and with no leadership from the federal government in 2020, we are underprepared at the state level for the SARS-CoV-2 epidemic as few individuals remain with knowledge from the early struggle against HIV. Specifically, the right data systems are not in place at most state DOHs to provide consistent and interpretable data streams allowing epidemiologists to make real-time assessments on epidemic progression and success of control efforts. State-level systems in the US require more funding from the federal government or centrally designed (and funded) reporting tools from the Centers from Disease Control and Prevention that would allow all states to consistently report the same high-quality data. The rationale for this systems upgrade would be to advance our surveillance systems to those of countries like New Zealand, Hong Kong, Singapore, Vietnam, Taiwan, and South Korea that have successfully controlled epidemic waves and introductions of SARS-CoV-2.

## 4 Methods

### 4.1 Case Data

Full description available in Supplementary Materials. Eleven data streams were assembled from three state DOH websites and data dashboards: (1) cumulative confirmed cases, (2) cumulative confirmed cases by age, (3) cumulative hospitalized cases, (4) cumulative hospitalized cases by age, (5) number of patients currently hospitalized, (6) number of patients in ICU currently, (7) number of patients on mechanical ventilation currently, (8) cumulative deaths, (9) cumulative deaths by age, (10) cumulative hospital deaths, (11) cumulative hospital discharges, with streams 6 and 11 missing in PA, and 10 and 11 missing in MA. Cumulative hospitalizations (data streams 3 and 4) in MA were reported as a subset of symptomatic cases in MA (via follow-up case investigations) and were excluded from the analysis. Reporting started on Feb 27 (RI), Mar 1 (MA), and Mar 6 (PA), and data sets used in this analysis use about 180 days of data through August 31. Age-specific counts often summed up to be less than the corresponding total daily counts of new symptomatic cases, new hospitalizations, or new deaths. This was common due to lack of age reporting in some proportion of cases. We assumed missing age-structured data to be missing completely at random when their sum was less than the total count. Data from random asymptomatic screening efforts (elderly care facilities, health care workers) were available for five months in RI and one month in MA. RI screening data were incorporated into the analysis to adjust the inference on the reporting fraction (*ρ*), as these individuals did not report to the health system but were sought out by the health system.

### 4.2 Mobility Data

The first set of mobility data is provided by the COVID-19 Mobility Network [19] and is derived from users of the Facebook mobile app with the location history option enabled, representing approximately 0.8% of MA, 1.1% of PA and 1.1% of RI of each state’s population. Each user’s location is binned into tiles, approximately 470m × 610m at Pennsylvania’s latitude. These are aggregated by home county and date, and reported as the fraction of users who remain in one tile for the whole day. In this paper, we report state-level data by weighting these proportions by each county’s population, per the U.S. Census’ 2019 estimates. These estimates are not adjusted for the demographics of Facebook’s user base.

The second set of mobility data is provided by social distancing metrics recorded by SafeGraph [59]. The data were derived from GPS pings of anonymous mobile devices. A common nighttime location leach device over a 6-week period was defined to be the device’s “home” and daily GPS pings were analyzed to determine whether the device exhibited certain behaviors including completely staying at home, working part time, working full time, etc. The counts were aggregated at the Census Block Group (CBG) level, which is the second-smallest geographical unit for which the US Census Bureau publishes data. A state-level percent at home fraction can be calculated by dividing the ‘completely at home’ devices in a state by the total devices in that state, however one step was taken prior to this calculation as outlined in the data analysis methodology for the Stay-At-Home Index provided by SafeGraph [60]. The step included was a correction for sampling bias at the CBG level by resampling with a stratified reweighting method described in the supplementary materials [61] (see Supplementary Materials Section 1.6.2).

### 4.3 Mathematical Model

A standard age-structured ordinary differential equations (ODE) model was used to describe the dynamics of SARS-CoV-2 spread in a single well-mixed population. The model includes 30 compartments for different clinical states including susceptible, exposed, asymptomatic, infected, hospitalized, in ICU, on mechanical ventilation. Multiple consecutive compartments are used for most clinical states to reduce the variance on length-of-stay in various stages in disease progression. Model diagram shown as Figure S2 and equations shown in the Supplementary Materials Section 2.

Model parameters fall into several categories including parameters on contact rates, lengths of stay in various clinical states, and probabilities of progression from one state to another. Daily community-level transmission-capable mixing rates *β*_*t*_ were inferred from the data, while age-specific contact rates for hospitalized individuals had to be fixed as too little data exist on these parameters during the March/April lockdown period. Asymptomatic individuals are assumed to be half as infectious as symptomatic individuals (similar to other models’ assumptions [62,63]).

Lengths of stay and age-specific probabilities of clinical progression were available from numerous data sets documenting COVID-19 hospitalized populations; details in Supplementary Materials Section 2, Table S1 and https://github.com/bonilab/public-covid19model. When certain clinical parameters were inferred, their median estimates were typically close to observed values in hospital or surveillance datasets. No data were available in RI, MA, or PA to infer the asymptomatic fraction for each age group, and these were obtained from a literature review (see Supplementary Materials Section 1.8); several specific hypotheses on asymptomatic transmission were compared with equal prior probabilities on age-specific symptoms models (Table 2); we ultimately used the Davies et al [25] fractions for the final model runs.

### 4.4 Statistical Inference

Given the various – and at times incomplete – data sources available for each state, we chose a flexible Poisson-Gamma process-based likelihood framework to facilitate inference of ODE model parameters while accounting for model uncertainty. In particular, the cumulative cases, hospitalizations, deaths, and hospital discharge data were assumed to be realizations of conditionally independent, inhomogeneous negative binomial processes, with time-varying process rates defined by the expected deterministic ODE output. The likelihood function for each age-structured data stream is then a product of independent, negative binomial increments, with means determined by the corresponding age-structured component of the ODE system over each increment. Means for observed new symptomatic cases were equal to ODE system predictions multiplied by a symptomatic reporting rate constant, and means for observed new hospitalized individuals were equal to ODE system predictions multiplied by a hospitalization reporting rate constant. When random screening data are available, we adjust the mean of the number of new symptomatics to include an additional additive term equal to the rate of random testing times the probability of a positive test. Dependence across data streams is assumed to be captured by the ODE system. Total data streams, summed over all age classes, were viewed as the sum of independent negative binomial random variables, and are as such negative binomial random variables themselves, with mean given by the sum of the age-specific means. When both age-structured and total data are available, we assume any missing age-structured data are missing completely at random, and approximate the joint likelihood of the total and age-structured counts by ignoring overdispersion and assuming that, conditioned on the total data, the age-structured counts are multinomially-distributed with probabilities proportional to the age-structured ODE means. Data on current hospitalizations, as well as current numbers in intensive care units, and current intubated individuals were modeled using daily increments. The daily change in intubated individuals, individuals in intensive care units (but not intubated) and hospitalized individuals (not in intensive care units) were each modeled as independent Gaussian random variables with means equal to the corresponding daily change predicted by the ODE system, and with unknown variances. Additional details on the likelihood framework can be found in Supplementary Materials, Section 3.

We chose a Bayesian approach to inference, allowing for appropriate penalization of time-varying parameters and a combination of strongly and weakly informative priors for parameters relating to clinical progression of disease. The composite rate parameter, *β*_*t*_, describing person-to-person contact mixing, is constructed via a cubic B-spline, with a random walk prior (penalized regression spline with 1^st^ order differences) on the B-spline coefficients to penalize overfitting. In RI and PA, the symptomatic reporting rate *ρ* is constructed as an I-spline with a similar prior; in MA it is assumed to be constant across time. Additional parameters found within the ODE system, including length of hospital stay and proportion of cases needing hospitalization within each age class are given uniform priors with bounds determined by expert judgement, while negative binomial dispersion parameters are given weakly informative exponential priors and Gaussian variance parameters are given conjugate inverse-gamma priors. Given these priors and the previously defined likelihood, we constructed a Markov chain Monte Carlo (MCMC) algorithm to sample from the posterior distribution of model parameters. Block updates for parameters were obtained using a random walk Metropolis-Hastings algorithm with an adaptive proposal distribution [64]. For each state, five independent chains were run for 300,000 iterations, with the first 100,000 samples discarded as burn-in. Convergence was assessed qualitatively across the five chains. R and C++ Code is posted at https://github.com/bonilab/public-covid19model.

## Data Availability

Posted on https://github.com/bonilab/public-covid19model

