## Supplementary Materials for "SARS-CoV-2 epidemic after social and economic reopening in three US states reveals shifts in age structure and clinical characteristics"

Date of this draft: November 16, 2020.

**5 Additional Figures** **25**

### 1 Data

Case counts of molecularly-confirmed SARS-CoV-2 (COVID-19) infections together with hospitalization counts from the same populations were obtained from three states – Rhode Island, Massachusetts, and Pennsylvania – from data posted by each state's Department of Health (DOH). As catchment populations and data definitions, formats, and completeness changed through time and varied by state, all data streams were cleaned and periodically validated for internal consistency. Discrepancies and changing patterns in epidemiology, care standards, passive surveillance/random screening, case definitions, and case clusters were discussed with DOH staff when possible.

#### 1.1 List of Data Streams

The eleven daily data streams considered were (1) cumulative confirmed cases, (2) cumulative confirmed cases by age, (3) cumulative hospitalized cases, (4) cumulative hospitalized cases by age, (5) number of patients currently hospitalized, (6) number of patients in ICU currently, (7) number of patients on mechanical ventilation currently, (8) cumulative deaths, (9) cumulative deaths by age, (10) cumulative hospital deaths, (11) cumulative hospital discharges.

#### 1.2 Rhode Island

Data for Rhode Island were tracked daily from the Rhode Island Department of Health (RIDOH) website reporting COVID-19 response data, which links to a publicly available datasheet that tracks all eleven data streams, with periodic interruptions to the availability of age-based data for new cases, hospitalizations, and deaths. Case data were originally reported with delays of one to five days, with most results being same day or one day delayed by the April reporting period. RIDOH staff on the data and epidemiology team periodically updated past results to move cases from 'day of reporting' to 'day of test', and all final and cleaned case data in Rhode Island were reported by 'day of test'.

#### 1.3 Massachusetts

Data from Massachusetts were obtained from the daily and weekly daily archive of the Massachusetts Department of Public Health's (MassDPH) case, testing, hospitalization, and death data. Confirmed daily case data were available by 'date of testing' (`CasesByDate.csv`), and age-structure of new cases was available (`Age.csv`, "Catchment 1") but only summed to a portion (typically 80% to 90%) of the total daily case numbers, a likely result of no age reporting in a number of hospitals. Confirmed daily death data (confirmed only, excluding probable) were available by 'date of death' (`DateOfDeath.csv`), and deaths by age were available (`Age.csv`) but again only summed to a portion (again, typically 80% to 90%) of the total daily case numbers (potentially the result of no age reporting from some locations).

Age-structured cumulative hospitalization were included in the data (`Age.csv`) but these had to be excluded as these hospitalization counts were obtained by following up with patients who had reported as symptomatic (thus, their denominator was symptomatic cases, not all infections). It was clear that this data stream was underreported as for a period in April current hospitalized exceeded cumulative hospitalized. Numbers of patients in ICU and on mechanical ventilation were available daily from April 4

(`Hospitalization from hospitals file.csv`).

Current hospitalization data for the April 7-14 date range were manually adjusted as a smaller set of hospitals was reporting for those days. After September 6, 2020, age-stratified cases, hospitalizations, and death are reported in MassDPH's weekly public health report (`weekly-covid-19-dashboard-data-9-30-2020.xlsx`) instead of the daily covid dashboard (`Age.csv`)

We assume that death data are not underreported. Current hospitalization data were obtained from 68 acute care hospitals (ACH) in Massachusetts, which represent all ACH in MA and would cover all or nearly all COVID-19 hospitalizations in MA.

Symptomatic case data were smoothed with a 7-day moving average to remove a weekly periodic signal (lower Sunday/Monday reporting).

##### 1.4 Pennsylvania

Daily new confirmed case data were pulled from the Pennsylvania Department of Health Coronavirus Update Archive. Counts are reported beginning with the first reported case on March 6. For the period from April 17 to May 21, confirmed cases were estimated from the reported values, which are noted to be the sum of confirmed and probable cases. Confirmed cases were estimated to be 97% of the combined total, based off of dates for which confirmed and probable counts were available separately, and match neighboring count data well. After May 21, daily confirmed counts (separate from probable cases) were made available. Beginning June 9, these counts were determined from the sum of confirmed cases reported at the county-level, as the state total was no longer reported on the archive. One outlier (April 16) was removed from the data set, as the daily change reported was inconsistent with neighboring counts. No data was available on June 8. For all dates, the 'confirmed only' data were used in data fitting.

Age groups for new cases from March 27 to June 7 were also available on the Pennsylvania DOH Archive. Age data were reported as the percentage of 'positive cases by age range to date' in the following age ranges: 0-4, 5-12, 13-18, 19-24, 25-49, 50-64, and 65+. These were translated into 10-year age bands by rebinning the counts under the assumption that the age probability density of case counts was relatively smooth. A monotonic spline was fit to the empirical cumulative distribution and then differentiated to estimate the 1-year age band (probability distribution function) of the original age categories. These were validated against known age breakdown data, and underreporting for the 80+ group was found, as would be expected for the 65+ age category. Beginning June 17, age-structured data was pulled data from Pennsylvania's online arcgis dashboard and scraped using the R package 'rvest'. Age data reported on the dashboard were binned in 10-year age groups, so no rebinning was necessary. For days when the age-structured case counts exactly matched the previous day's counts (apparently because they had not been updated), the data were omitted.

Daily death counts are reported on Pennsylvania's arcgis dashboard as well, beginning with the first reported death on March 18. These numbers are retrospectively updated to reflect the date of death. According to the dashboard: "Death data is based on the date of death as reported to EDRS. Death counts for prior dates will change as additional death records are registered or amended." Beginning May 17, age-structured death counts (binned into 5-year age groups) were available from weekly reports through the Pennsylvania DOH Coronavirus Archive. These reports also included the breakdown of the location of deaths (i.e. hospital, hospice, long-term living facilities, and residence). More frequent age-structured death data were collected from daily pulls from the Pennsylvania arcgis dashboard beginning June 17.

Cumulative hospitalization counts were available on the Pennsylvania DOH Archive from March 27 to April 6. From April 17 to May 21, these counts were calculated as the sum of the approximated age-structured hospitalization counts. These age-structured values were estimated from the reported percentages of 'hospitalizations by age range to date', available for this time period on the Pennsylvania DOH Archive. Hospitalization counts are not reported after May 21.

The currently hospitalized population in Pennsylvania has been reported on the online arcgis dashboard since April 17. As in MA, a problem with this data stream is the fact that in mid-April, the current hospitalization counts exceed the cumulative hospitalization counts reported, suggesting that 'current' numbers are drawn from a different (larger) set of hospitals than the 'cumulative' numbers. For this reason, a 'hospitalization reporting fraction' was included in the PA analysis to account for the fact that only a portion of hospitals in PA are/were reporting new hospitalizations to the PA DOH.

The current number of patients in the intensive care unit is not reported for Pennsylvania. Current numbers of patients on ventilator are reported on the online dashboard, alongside the current hospitalized number, and have been available since April 17. Cumulative hospital discharges are not available.

#### 1.5 Data on testing and cases in elderly care facilities

Data on testing and cases in elderly care facilities were obtained for Rhode Island and Massachusetts from the same sources as described in Sections 1.2 and 1.3, respectively. The extractions of the partial time series below were not sufficient for some of our planned analyses, and only data from Table S2 were used to correct for the amount of testing done in asymptomatic populations.

##### 1.5.1 Rhode Island

First, weekly cumulative data by long term care and assisted living facilities were extracted from the tables with the prefix '`RI_Cumulative_Long_Term_Care_and_Assisted_Living_`'. Each table, corresponding to a single day, was summarized and five categories of information were obtained: the date of the table (i.e., the version), the date to which the values in the table correspond, the number of facilities included, the cumulative number of cases, and the cumulative number of resident fatalities. Cases and fatalities were described using strings defining intervals (e.g., '15 to 19' or 'Fewer than five'); first the numerical endpoints were extracted, and then three types of sums calculated: of lower endpoints, midpoints and upper endpoints, respectively. Since all tables had been corrected multiple times, resulting in duplicate records, only the latest versions for every reporting day were used in the subsequent analysis. This resulted in eight data points ranging from April 29 to June 11. The number of facilities included increased from 44 to 51 over the time range; this was due to the fact that only facilities with at least two cases were included.

Second, facility fatalities by location of death were obtained from the tables with the prefix '`RI_Copy_of_Facility_Fatalities_by_Location_of_Death_`'. These were provided as proportions corresponding to three types of facilities: hospices, hospitals, and long term care facilities. Here, we obtained 40 data points ranging from May 12 to June 19, with only minor changes in individual values over time: the hospice proportion decreased from 9% to 8%, the long term care facilities proportion was monotonically increasing from 60% to 62%, and the proportion of hospital fatalities fluctuated between 29% and 32%.

###### 1.5.2 Massachusetts

External dashboard Excel files were identified in the raw data archives as files matching '`Ext*.xlsx`'. This resulted in 33 external dashboards providing data on nursing homes from April 29 to May 31. From each dashboard, the following types of information were extracted (if available): the date, the cumulative number of tests in nursing homes, the cumulative number of cases in nursing homes, and the cumulative number of deaths. Dates were parsed from the filenames. The cumulative number of tests and cases were obtained from the '`Nursing home testing`' table, which was first converted to a textual representation, cropped (to remove variable-length header), converted back to a table format, which was then used to extract the "`Yesterday`" column; the numbers in the column were then sorted and two highest numbers summed up and reported, corresponding to the total tests completed in the EMS and mobile setting, respectively. If any of the values was "TBD - waiting on data", 0 was used instead. The cumulative number of deaths was calculated from the table called 'Nursing home' in a similar fashion; nevertheless, these data were present only for May 27.

##### 1.6 Mobility Data

Both Facebook and SafeGraph apply differential privacy to the provided data by adding Laplacian noise to stay-at-home counts and discarding information for locations with too few users or devices [1][13].

Both sources are limited by their observed populations, which are not representative of states in question. Except for re-weighting by the total population of reported counties/CBGs, no adjustments are made to estimate mobility for the general public, or to make the two metrics comparable.

###### 1.6.1 COVID-19 Mobility Network

The COVID-19 Mobility network provides access to data from Facebook's Data for Good program, specifically its Movement Range Trends dataset. (Note: This data has since been made publicly available.) This is derived from the tracks of Facebook app users with the 'location history' option enabled, which amounts to approximately 1% of the population in each state studied. Location is binned into tiles approximately $470m \times 610m$ at Pennsylvania's latitude. A user is counted if their location is reported in at least three different hours of a given day. They are counted as 'staying put' if all their reported locations are in a single tile.

These data are provided by Facebook's 'GeoInsights Portal' in the form of daily csv files (`Movement Range_CH_NUTS3_YYYY-MM-DD.csv`) with one US county per line. From these we extract the columns `ds`, `all_day_ratio_single_tile_users`, and `external_polygon_id` corresponding respectively to the date, stay-home fraction and county FIPS code, respectively.

We create state-level aggregations by weighting these data by U.S. Census Bureau 2019 county population estimates (co-est2019-alldata.csv). (Note: Facebook uses the old FIPS code for Oglala Lakota County, which changed from 46113 to 46102 in 2015.)

###### 1.6.2 SafeGraph

Census Block Groups (CBGs) are the second-smallest statistical area reported by the U.S. Census Bureau, containing about 600–3000 people each [6]. A sampling bias is introduced in the SafeGraph mobility data when CBGs are not all sampled at equal rates relative to their true populations. For example, urban areas may be oversampled due to a greater usage rate of smart phones compared to rural areas. Fortunately, the true populations of CBGs are known from the official US Census Bureau. When the true population fraction of a CBG in a state is compared to the fraction of SafeGraph observations of that CBG in the state, we can identify whether the CBG is over or underrepresented in the samples. Stratified reweighting allows us to increase the weight of underrepresented CBGs and decrease the weight of overrepresented CBGs . A reweighting factor is therefore necessary and calculated by using a stratification method as reported in the equation below.

$$W_{CBG,S} = \frac{N_{CBG}}{N_S} \frac{n_S}{n_{CBG}} \tag{S1}$$

In the above equation, $W_{CBG,S}$ represents the weighting factor for a CBG in state $S$. $N_{CBG}$ represents the true population of the CBG as reported in the US Census Bureau, $N_S$ represents the true population of $S$ as reported by the Census Bureau, $n_{CBG}$ represents the number of SafeGraph observations sampled from the CBG and $n_S$ represents the total number of SafeGraph observations sampled from $S$. From this equation, it is true that as the sample fraction of CBG in $S$ approaches the true population fraction of CBG in $S$, $W$ gets closer to one. Within a given state $S$, $W_{CBG,S}$ is greater than one for CBGs which are under-represented in the SafeGraph data, and vice-versa. The raw data for a CBG is multiplied by these weighting factors to correct for sampling bias.

#### 1.7 Variable Names

Our eleven data streams are coded with the following variable names:

$$\begin{array}{rl} x_{a,t} & \text{cumulative case counts by age group } a \\ y_{a,t} & \text{cumulative hospitalizations by age group } a \\ z_{a,t} & \text{cumulative hospital deaths by age group } a \\ w_t & \text{cumulative out-of-hospital deaths (not available by age group)} \\ u_t & \text{cumulative hospital discharges (not available by age group)} \end{array} \tag{S2}$$

and

$$\begin{array}{rl} h_t & \text{current number of hospitalized individuals at time } t \text{ (includes } c \text{ and } v \text{ below)} \\ c_t & \text{current number of individuals in the ICU at time } t \text{ (includes } v \text{ below)} \\ v_t & \text{current number of individuals on a ventilator at time } t \end{array} \tag{S3}$$

For some states and some time periods, age-structured data are not available, but age-aggregated data are available. This is common for cumulative case counts and cumulative hospitalizations and can occur for all variables; in some cases, age-stratified data are reported weekly or irregularly. We refer to these aggregated data as

$$
\begin{aligned}
x_t = &\quad \text{cumulative new case counts (all ages)} \\
y_t = &\quad \text{cumulative new hospitalizations (all ages)} \\
z_t = &\quad \text{cumulative new deaths (all ages)}
\end{aligned}
\tag{S4}
$$

Age-structured counts of cumulative cases, hospitalizations, and deaths can be available at the same time as the corresponding cumulative counts $x_t$, $y_t$, and $z_t$, and it is common for the summed age-structured counts to record fewer individuals than the reported total counts. That is,

$$x_t > \sum_a x_{a,t} \tag{S5}$$

is a common situation due to missing age information for some patients; in this case we assume that the age data are missing completely at random. There are also occurrences where the age-sums are larger than the reported totals, and this occurs occasionally because the cumulative age data have been put in the incorrect row in a data base (e.g. one week earlier than where they should be). These latter cases were handled individually, in order to not assume that one data stream is correct while the other is not, and verified with a state DOH when possible.

#### 1.8 Asymptomatic fraction by age

To assess the percentage of SARS-CoV-2 infections that never progress to symptoms, literature review was conducted during the course of our analysis to identify studies that reported asymptomatic fractions by age group. A total of 12 studies/datasets were included: 8 peer-reviewed articles, 2 CDC MMRW reports, 1 pre-print, and 1 news report. Four of these studies report asymptomatic infection that was confirmed, with follow-up, to remain asymptomatic [16, 19, 26]; these are shown with yellow boxes/circles in Figure S1. Pham et al. described cases observed during Vietnam's quarantine and isolation effort during the first 100 days of the pandemic. In this paper, confirmed infections that never developed symptoms during the 14-day quarantine are defined as asymptomatic infection. Kimball et al. described infections among the residents from a skilled nursing facility in King County, Washington state. Follow-up assessment was conducted, and the infected individuals that never showed symptoms were defined as asymptomatic. Van Vinh Chau et al. described SARS-CoV-2 infections in a hospital designated for COVID-19 patients in Vietnam. Infected individuals that never developed symptoms during a 14-day quarantine are classified as asymptomatic. Lytras et al. described confirmed cases among passengers on repatriation flights in European countries; infected passangers that did not report symptoms for the duration of the study (17 days) were defined as asymptomatic. Other studies Bi et al. [4], Breslin et al. [5], Cereda et al. [7], Imperial College COVID-19 Response Team et al. [14], Kim et al. [15], Mizumoto et al. [20], Payne et al. [22], So and Smith [25] did not contain sufficient follow-up to ensure that patients were truly asymptomatic, thus it is possible that some of these studies may include pre-symptomatic individuals; these are shown with gray boxes/circles in Figure S1

Combining the four studies that include asymptomatic cases only, the weighted-mean of the asymptomatic fraction across all ages is 43.6%. The asymptomatic fraction by age is: [10-19] : 38.1%; [20-29]: 65.7% ; [30-39]: 43.3%; [50-59]: 43.1%; [60-69]: 40.9%; 80+: 13%. There is insufficient data to infer the asymptomatic fraction in the age intervals [0-9], [40-49], [70-79]. This is either because we excluded the studies that were clinical reports due to potential bias, or the sample size in the studies is too small to draw a conclusion. Grouping into 20-year age bands, the asymptomatic fractions are [0-19] : 38.1%; [20-39]: 46.0%; [40-59]: 43.1%; [60-79]: 40.9%; [80+]: 13.0%.

The five probabilities above ("simple average" in Table 2 of the main text) are compared to five other asymptomatic models in our analysis.

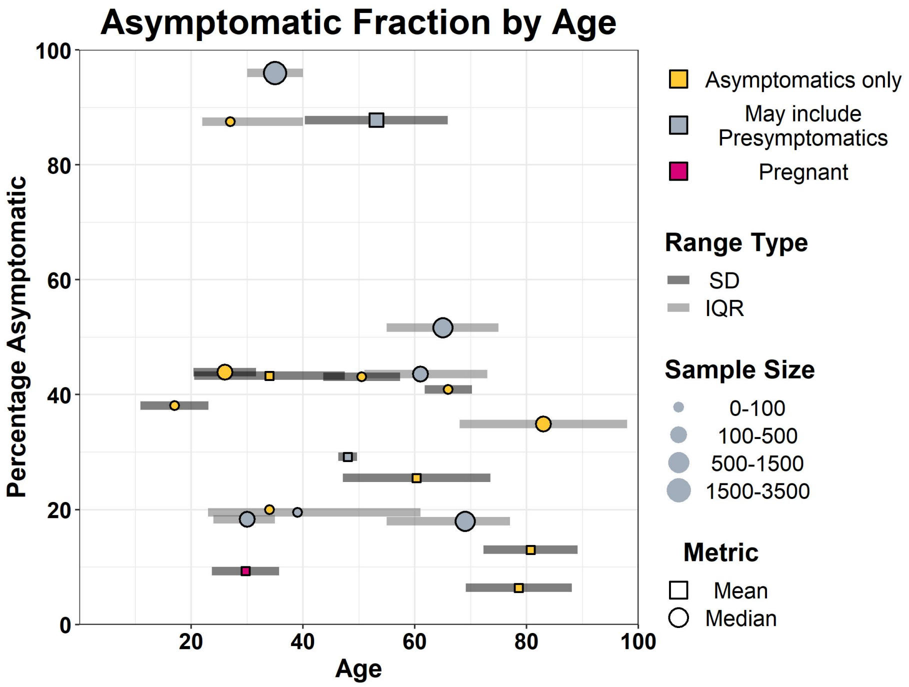

Figure S1: Asymptomatic Fraction by Age

Boxes (means) and circles (medians) show the percentage of SARS-CoV-2 infections that are asymptomatic for a particular age group. Yellow markers correspond to studies that included confirmed asymptomatics (with sufficient follow-up) while gray markers correspond to studies that may have included pre-symptomatic individuals. One study (magenta) looked at pregnant women only. Gray bars indicate either the standard deviation in age or the interquartile range of age for the selected group. Larger markers correspond to larger sample sizes.

#### 2 Mathematical Transmission Model

A classical ordinary differential equations (ODE) model was used to model infection and clinical progression of SARS-CoV-2, with compartmental diagram shown in Figure S2. The model is age-structured with nine age classes broken into 10-year age bands, with $\geq 80$ as the last age class. Age-specific forces of infection ($\Lambda_a$) determine how quickly individuals move from $S_a$ (susceptibles in age class $a$) to $E_{1,a}$ (stage 1 of the exposed class for age class $a$). All other transitions in the model are linear. Classes with a circled number in the upper right are broken up into that many stages. For example, the exposed state is six stages long ($E_{1,a}$ to $E_{6,a}$) with each stage being one day long. The mean duration of the exposed period is six days, and the coefficient of variation of this duration is $1/\sqrt{6} = 0.41$.

The classes in the compartmental model are susceptible ($S$), exposed ($E$), infected and asymptomatic ($A$), infected and symptomatic but not hospitalized ($I$), hospitalized during the acute-phase of the infection on the medical-floor level of care prior to potential ICU admission ($H_A$), admitted to the intensive care unit (ICU) after a recent hospital admission ($C_A$), on a mechanical ventilator in the ICU (V), removed from mechanical ventilation but still in the ICU ($C_R$), discharged from ICU and on the medical-floor level of care in the hospital ($H_R$), recovered from a non-hospitalized infection ($R$), and discharged from the hospital ($R_{\text{HOSP}}$). The 'A' subscript indicates acute phase, and the 'R' subscript is meant to indicate that the patient is recovering.

Figure S2 shows all the transitions in patient clinical progression, including death which is shown with dashed lines. Transition probabilities are explicitly included as model parameters and are not approximated as ratios of exit rates from classes.

The force of infection on age class $a$ is

$$\begin{aligned}
\Lambda_a \quad &= \quad \beta_t \cdot \left[ \sum_{b=1}^{9} \sum_{j=1}^{6} c_{a,b} \varphi_E E_{j,b} \; + \; \sum_{b=1}^{9} \sum_{j=1}^{4} c_{a,b} \varphi_A A_{j,b} \; + \; \sum_{b=1}^{9} \sum_{j=1}^{4} c_{a,b} I_{j,b} \right] \\
&+ \quad \beta_{\text{HOSP}} \sum_{b=1}^{9} \sum_{j=1}^{4} c^H_{a,b} (\varphi_H H_{A,j,b} + \varphi_{H_R} H_{R,j,b}) \\
&+ \quad \beta_{\text{CRIT}} \sum_{b=1}^{9} \sum_{j=1}^{1} c^C_{a,b} (\varphi_C C_{A,j,b} + \varphi_C C_{R,j,b}) \\
&+ \quad \beta_{\text{VENT}} \sum_{b=1}^{9} \sum_{j=1}^{6} c^V_{a,b} \varphi_V V_{A,j,b} \qquad \text{(S6)}
\end{aligned}$$

where $b$ runs over age classes and $j$ runs over stages of a class.

The parameter $c_{a,b}$ is the contact rate between age groups $a$ and $b$, which is symmetric in our model and simply the product of the mixing rates of the two age classes. Contact rates for hospitalized individuals are chosen so that children and teens do not visit hospitalized patients, $\geq 70$ individuals visit hospitalized patients with a relative contact parameter of 0.2 (no contact or visits except for EOL circumstances), and 20–69 individuals have a relative contact parameter of 1.0 as hospital and medical staff. $\beta_{\text{HOSP}}$ is chosen to be 0.2 where $\beta = 1.0$ is the level of population mixing during March 5-15, before the initial lockdowns began. No visits to the ICU are allowed. Patients in the ICU only have contact with medical/hospital staff, aged 20–69. $\beta_{\text{ICU}} = \beta_{\text{VENT}} = 0.2$.

The $\varphi$ parameters represent the relative infectivity of certain compartments with $\varphi = 1.0$ for non-hospitalized symptomatic individuals (first two stages only $I_1$ and $I_2$). We set $\varphi_A = \varphi_E = 0.5$, $\varphi_{I_3,I_4} = 0.75$ in the second half of the symptomatic stage. $\varphi_H = 1.0$ in the acute stage of hospitalization and $\varphi_{H_R} = 0.2$ in the convalescent stage of hospitalization. $\varphi_C = 1.0$ and $\varphi_V = 0.1$. These values are largely unknown and they were set based on common assumptions in the literature and conversations with attending clinicians.

The parameter $\beta_t$ is a time-dependent population-mixing parameter, whose values are inferred and constrained by a spline structure (see Equation S20).

Admission to the ICU, if it occurs, typically occurs early in the hospitalization period. Patients can be admitted immediately after presentation to the emergency department or after a short medical-floor hospital stay [24]. Thus, individuals can enter $C_A$ from $H_{A,1}$ but not $H_{A,2}$ through $H_{A,4}$. Time on ventilator is broken up into six stages, as death occurs more quickly than recovery. Total time on mechanical ventilation is a parameter that is inferred with a starting value of and constrained to be close to 10.8 days [3, 8, 12, 29]. Dashed lines in the class diagram indicate death. Individuals can die either in the ICU or on the medical-floor level of care. Non-hospitalized individuals can die at home, and this transition is included mainly to model death in congregate care facilities (nursing homes) where individuals with severe COVID-19 infections are not counted as hospitalized. These individuals are mostly $\geq 70$ and in Rhode Island there is sufficient data to estimate a 'death-prob-home' parameter for the 60-69 age-group as well.

Dynamical equations for the susceptible classes are

$$\dot{S}_a \;=\; -\sigma_a \, \frac{\Lambda_a}{N} \, S_a \tag{S7}$$

where $\sigma_a$ is the relative susceptibility of age-class $a$ (set to 0.6 for $< 20$ and 1.0 for everyone else ) and $N$ is the population size. Other parameters not described above are listed in Table S1. All other differential equations are sums of linear terms with transition rates determined by 'lengths of stay' in each class (above and in Table S1), and probabilities determining whether someone progress to (for example) death, a milder clinical state, a more severe clinical state, or the same clinical state.

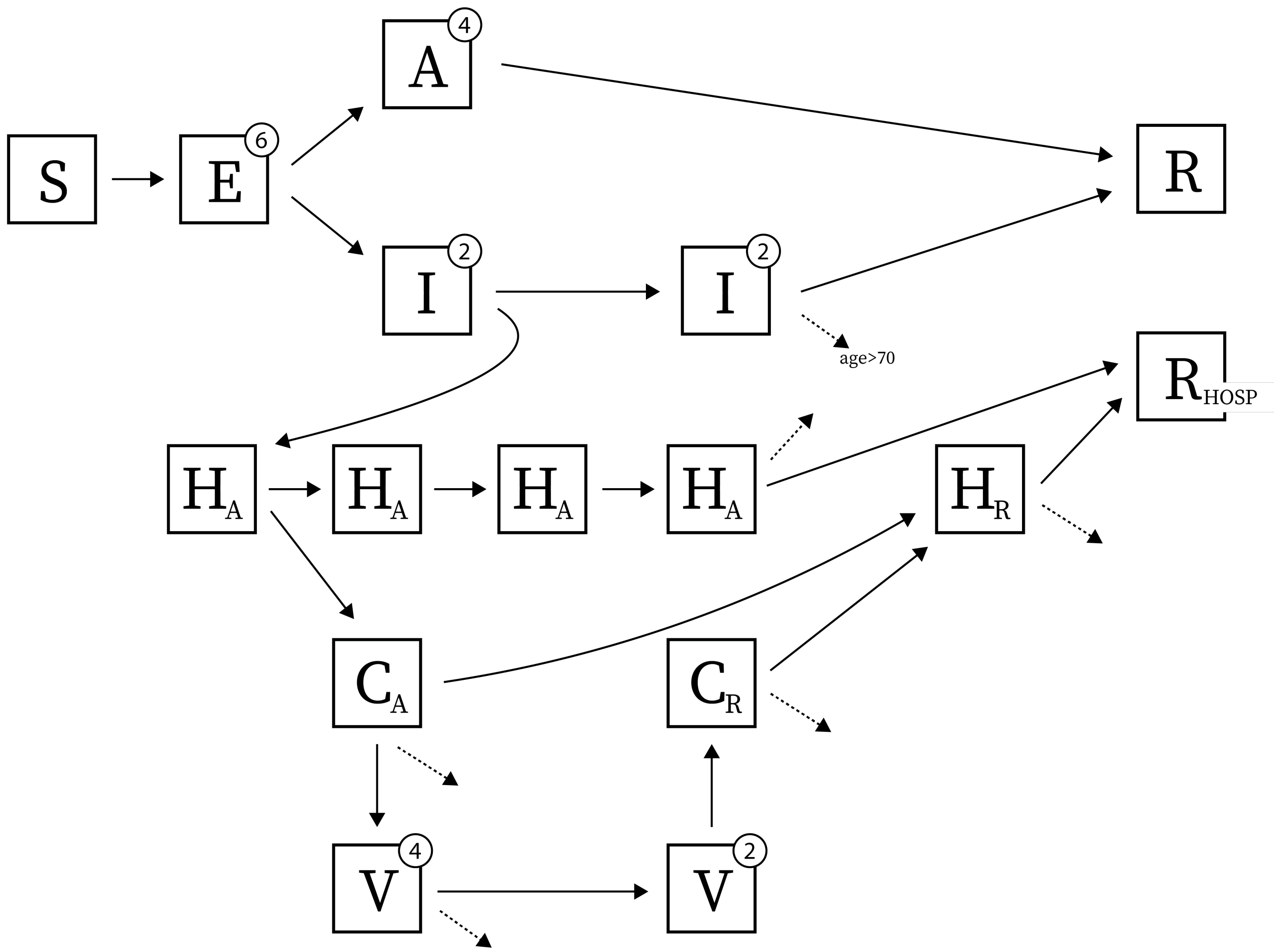

Figure S2: **Class Diagram of Mathematical Transmission Model**

A circle in the top-right corner of a class indicates that the class is broken up into $n$ consecutive stages, to reduce the variance of the length-of-stay in that class. Arrows leaving classes only leave the last stage of that class. For this reason, the four stages of the acute-phase hospitalized class $H_A$ are drawn individually so that it's clear that progression to the ICU (class $C_A$) occurs from stage $H_{A,1}$ and not the other stages. Dashed arrows indicate death. Non-hospitalized individuals in $I_4$ can die if they are in the older age classes, $\geq 70$ for MA and PA and $\geq 60$ for RI (this models death at congregate care facilities). Time-on-ventilator (class $V$) is longer for survivors than for non-survivors and this is modeled by a 6-stage length-of-stay with individuals dying after stage 4 and progressing off mechanical ventilation after stage 6. All transitions in the model are linear, except the transitions from $S_a$ to $E_{1,a}$ which are determined by the age-specific force of infection $\Lambda_a$.

Table S1: Parameter values in transmission model.

| parameter | value |
|---|---|
| mean duration of exposed period | 6.0 days |
| mean duration of asymptomatic infection | 5.3 days (no data on this) |
| mean duration in $I_1$ and $I_2$, i.e. the time from symptoms to hospitalization | between 2 and 5 days (fit) |
| mean duration in $I_3$ and $I_4$ | 7.0 days (fixed) |
| mean duration in $H_1$ through $H_4$, i.e. mean duration of medical-floor hospital stay | prior distribution centered around 10.7 days [17] |
| mean time in $C_A$ | 2.0 days |
| mean time in $C_R$ | 2.6 days [3] |
| mean time in $H_R$ | 2.5 days [no source, discussion with clinicians] |
| mean time on ventilator (survivors) | prior distribution centered around 10.8 days [3, 8, 12, 29] |
| probability of asymptomatic infection by age | 0.71 (0-9 age group), 0.79 (10-19), 0.73 (20-29), 0.67 (30-39), 0.60 (40-49), 0.51 (50-59), 0.37 (60-69), 0.31 (70-79), 0.31 (80+); [9] and see section 1.8 |
| probability of hospitalization by age | fit for RI and PA; fitted RI values used for MA |
| probability of death at home | zero for $< 60$; fit for $\geq 60$ |
| probability of progression from hospital to ICU, by age | starting values in model fit are: 0.304 (0-9 age group), 0.293 (10-19), 0.2825 (20-29), 0.301 (30-39), 0.463 (40-49), 0.4245 (50-59), 0.460 (60-69) 0.4835 (70-79), 0.416 (80+) [17]; a single multiplicative scaling factor was fit for each state and for each of two epidemic periods. |
| probability of progression from ICU to ventilation | prior distribution centered around 0.75 |
| probability of death while on ventilator, by age | 0.031 (0-9 age group), 0.051 (10-19), 0.15 (20-39) $0.4 \cdot v$ (40-49), $0.46 \cdot v$ (50-59), $0.585 \cdot v$ (60-69) 0.7 (70-79), 0.9 (80+) [12, 28]; parameter $v$ is fit as there is substantial variation in the 40-70 age group for this parameter. |
| probability of non-ICU hospital death | 0.025 (70-79 age group), 0.050 (80+ age group); [no source] |
| probability of death after being removed from mechanical ventilation | 0.0945 (60-79 age group), 0.189 (70-79 age group), 0.378 (80+ age group) [12] |

### 3 Likelihood and Inference

#### 3.1 Daily New Count Data

Daily new case counts are assumed to follow a negative binomial distribution with dispersion parameter $k_1$. The expected mean number of new symptomatic cases in each age group, $\rho_t \cdot \tilde{J}_{a,t}(\theta)$, is obtained from the ODE model, with $\theta$ representing the parameters that are input into the ODEs (i.e., contact rate, length of hospital stay, etc). The parameter $\rho_t$ is the fraction of symptomatic cases that are expected to be observed by the health system; $\rho_t$ is the reporting rate at time $t$. The $\sim$ symbol is used to indicate first differencing across time steps, and

$$\tilde{J}_t \;=\; J_t \;-\; J_{t-1}$$

is the daily incidence while the $J_t$-variables are the cumulative incidence. Differencing may be done across multiple time steps if data are missing for certain time steps.

For daily new confirmed case data, the likelihood of these data is treated differently depending on whether age-data exist or not. Let $\mathbf{T} = \{1, 2, \ldots, t_f\}$ be the set of all times for which data exist. And, let $\mathbf{T}_a$ and $\mathbf{T}_m$ be a partition of $\mathbf{T}$, where age data are present ($\mathbf{T}_a$) and age data are missing ($\mathbf{T}_m$). $\mathbf{T} = \mathbf{T}_a \cup \mathbf{T}_m$. For convenience, we treat these two disjoint subsets as the ordered tuples:

$$\begin{aligned} \mathbf{T}_a &= (t_1, t_2, t_3, \ldots, t_n) \\ \mathbf{T}_m &= (t_1^*, t_2^*, t_3^*, \ldots, t_m^*) \end{aligned}$$

When age data exist, in each age group we calculate the likelihood of new cases since the last time point that age data were available. For example, on the second day that age data are available, we write the likelihood as

$$L_1(\, x_{t_2} - x_{t_1} \,,\, x_{a,t_2} - x_{a,t_1} \,) \;=\; P(\, x_{t_2} - x_{t_1} \,) \cdot P(\, x_{a,t_2} - x_{a,t_1} \mid x_{t_2} - x_{t_1} \,) \tag{S8}$$

where the left-hand side above is a shorthand for the joint likelihood of new total cases and new cases in each of the eight age groups. Again, using $\sim$ as a shorthand for first differencing across time steps, we can re-write the above equation compactly as

$$\begin{aligned} L_1(\, \tilde{x}_{t_2} \,,\, \tilde{x}_{0,t_2} \,, \ldots ,\, \tilde{x}_{8,t_2}) &= P(\, \tilde{x}_{t_2} \,) \cdot P(\, \tilde{x}_{0,t_2} \,,\, \tilde{x}_{1,t_2} \,,\, \ldots \,,\, \tilde{x}_{8,t_2} \mid \tilde{x}_{t_2} \,) \\ &= \text{NegBin}(\, \tilde{x}_{t_2} \,;\, \rho_{t_2}\tilde{J}_{t_2}, k_1 \,) \cdot \text{MultiNom}(\, \tilde{x}_{0,t_2} \,,\, \ldots \,,\, \tilde{x}_{8,t_2} \,;\, \tilde{x}_{t_2}, \{\tilde{J}_{a,t_2}/\tilde{J}_{t_2}\} \,) \end{aligned} \tag{S9}$$

with the parameters of the probability mass functions above written after the semi-colon. The parameter $k_1$ is the dispersion parameter for the negative binomial distribution and $\rho_{t_2}\tilde{J}_{t_2}$ is the mean. The negative-binomial parameterization used is

$$P(X = \tilde{x}_{t_j}) \;=\; \frac{\Gamma(\tilde{x}_{t_j} + k_1)}{(\tilde{x}_{t_j})! \,\cdot\, \Gamma(k_1)} \left( \frac{k_1}{\rho_{t_j}\tilde{J}_{t_j} + k_1} \right)^{k_1} \left( \frac{\rho_{t_j}\tilde{J}_{t_j}}{\rho_{t_j}\tilde{J}_{t_j} + k_1} \right)^{\tilde{x}_{t_j}} ,$$

from which the variance can be written as $\rho_{t_j}\tilde{J}_{t_j} + (\rho_{t_j}\tilde{J}_{t_j})^2/k_1$. Our model for the age data, conditioned on the total data, is multinomially distributed, with probabilities proportional to the ODE-derived expected age-structured incidence. This is the likelihood that would result if each age-structured data stream were independent of each other, and each were Poisson distributed.

If age data do not exist for a time point, we write

$$L_2(\ x_{t_j^*} - x_{t_j^*-1}\ ) \ = \ \text{NegBin}(\ x_{t_j^*} - x_{t_j^*-1}\ ;\ \rho_{t_j^*} \tilde{J}_{t_j^*}, k_1\ )\ , \tag{S10}$$

and note that the first differencing above is done across exactly one time step, meaning that the time step $t_j^* - 1$ could have age data. The likelihood of all of the reported data on symptomatic confirmed cases is then

$$\prod_{j=2}^{n} L_1(\ \tilde{x}_{t_j}\ ,\ \tilde{x}_{0,t_j}\ , \ldots,\ \tilde{x}_{8,t_j}) \ \cdot\ \prod_{j=1}^{m} L_2(\ \tilde{x}_{t_j^*})\ \ . \tag{S11}$$

#### 3.2 Daily New Hospitalization Data

New hospitalization data (i.e. daily incidence of hospitalization) are treated identically to the symptomatic confirmed case data. A hospital reporting parameter $\rho_H$ is used to indicate that in PA and MA, the new daily hospitalization numbers appear to be incomplete. The dispersion parameter for the daily reported numbers is $k_2$. As before, time points are separated into those that have age data ($a$) and those are missing ($m$) age data:

$$\begin{aligned} \mathbf{T}'_a &= (t_1, t_2, t_3, \ldots, t_n) \\ \mathbf{T}'_m &= (t_1^*, t_2^*, t_3^*, \ldots, t_m) \end{aligned}$$

Using the same negative binomial and multinomial approach as for the case data, the likelihood for the hospitalization incidence data stream can be written as

$$\prod_{j=2}^{n} L_3(\ \tilde{y}_{t_j}\ ,\ \tilde{y}_{0,t_j}\ , \ldots,\ \tilde{y}_{8,t_j}) \ \cdot\ \prod_{j=1}^{m} L_4(\ \tilde{y}_{t_j^*})\ \ . \tag{S12}$$

#### 3.3 Daily New Death Data

Death data can be broken down by home and hospital deaths, and they can also be broken down by age. Again, the age data may only appear at certain time steps, but the home and hospital death data streams typically have complete day to day reporting. The ODE model has state variables for home deaths ($D_{\text{HM}}$) and hospital deaths ($D_{\text{HP}}$), and we use $D = D_{\text{HM}} + D_{\text{HP}}$ for the total death count. As before we break the time points into those that have age data ($a$) and those are missing ($m$) age data:

$$\begin{aligned} \mathbf{T}''_a &= (t_1, t_2, t_3, \ldots, t_n) \\ \mathbf{T}''_m &= (t_1^*, t_2^*, t_3^*, \ldots, t_m) \end{aligned}$$

When age data are present, the joint probability of the age, home death, and hospital death data can be separated like so

$$P(\text{home deaths, hospital deaths, age-stratified deaths}) = P(\text{age}|\text{hosp}, \text{home}) \cdot P(\text{home}) \cdot P(\text{hosp})$$

since the at-home deaths and the hospital deaths are independent. We write the likelihood as

$$
\begin{aligned}
L_5(\, w_{t_2} - w_{t_1} \,,\, z_{t_2} - z_{t_1} \,,\, z_{a,t_2} - z_{a,t_1} \,) \;&=\; P(\,(z_{t_2} - w_{t_2}) - (z_{t_1} - w_{t_1})\,) \\
&\times\; P(\, w_{t_2} - w_{t_1} \,) \\
&\times\; P(\, z_{a,t_2} - z_{a,t_1} \mid \text{total deaths}\,) \,.
\end{aligned}
\tag{S13}
$$

For any timestep $j$, this can be written more compactly as

$$
\begin{aligned}
L_5(\,\tilde{w}_{t_j} \,,\, \tilde{z}_{t_j} \,,\, \tilde{z}_{0,t_j} \ldots \tilde{z}_{8,t_j}) \;&=\; P(\,\tilde{z}_{t_j} - \tilde{w}_{t_j}\,) \cdot P(\,\tilde{w}_{t_j}\,) \cdot P(\,\tilde{z}_{a,t_j} \mid \text{total deaths}\,) \\
&=\; \text{NegBin}(\,\tilde{z}_{t_j} - \tilde{w}_{t_j} \,;\, \tilde{D}_{\text{HP},t_j}, k_3\,) \cdot \text{NegBin}(\,\tilde{w}_{t_j} \,;\, \tilde{D}_{\text{HM},t_j}, k_4\,) \cdot \\
&\quad \cdots \text{MultiNom}(\,\tilde{z}_{0,t_j} \cdots \tilde{z}_{8,t_j} \,;\, \tilde{z}_{t_j} \,,\, \{\tilde{D}_{a,t_j}/\tilde{D}_{t_j}\}\,) \,.
\end{aligned}
\tag{S14}
$$

When age data are not available this is

$$
L_6(\,\tilde{w}_{t_j^*} \,,\, \tilde{z}_{t_j^*}\,) \;=\; \text{NegBin}(\,\tilde{z}_{t_j^*} - \tilde{w}_{t_j^*} \,;\, \tilde{D}_{\text{HP},t_j^*}, k_3\,) \cdot \text{NegBin}(\,\tilde{w}_{t_j^*} \,;\, \tilde{D}_{\text{HM},t_j^*}, k_4\,) \,,
$$

and the likelihood across all time points is

$$
\prod_{j=2}^{n} L_5(\,\tilde{w}_{t_j} \,,\, \tilde{z}_{t_j} \,,\, \tilde{z}_{0,t_j} \ldots \tilde{z}_{8,t_j}) \;\cdot\; \prod_{j=1}^{m} L_6(\,\tilde{w}_{t_j^*} \,,\, \tilde{z}_{t_j^*}) \;. \tag{S15}
$$

#### 3.4 Data on Current Number of Patients in Hospital, ICU, and on Ventilators

The model tracks current hospitalizations ($H_t$, summed across age classes), current ICU occupancy ($C_t$), and current number of patients using ventilators ($V_t$). Note that in the model these are disjoint groups, so $H_t$ represents hospitalized patients that are not in the ICU and $C_t$ represents ICU patients that are not on ventilators. The data on the other hand are typically nested so that $h_t$ includes all hospitalized patients including ICU patients $c_t$ and patients on ventilators $v_t$. The likelihood equations for the 'current' data streams are written down as normal distributions around the first differences of the hospitalized, ICU, and ventilated populations. If first differencing cannot be done because of missing data, time points are simply excluded. We assume independent Gaussian likelihoods for the increments of observed quantities corresponding to $H_t$, $C_t$, and $V_t$, and model the observations of sums of these using standard multivariate normal theory.

The ventilated patient counts are thus modeled as:

$$
L_7(\,\{v_t\}\,) = \prod_{t=2}^{t_f} \varphi_{\sigma_v}(\,\tilde{V}_t \;-\; \tilde{v}_t\,)
$$

where $\varphi_{\sigma_v}$ is a normal PDF with mean zero and standard deviation $\sigma_v$. The likelihoods for the data streams of the ICU patient counts is calculated by subtracting out the ventilated patients

$$L_8(\ \{c_t\}\ ) = \prod_{t=2}^{t_f} \varphi_{\sigma_c}(\ \tilde{C}_t\ -\ (\tilde{c}_t - v_t)\ )$$

and the model for the hospitalization counts are similarly constructed by subtracting the ICU patient counts

$$L_9(\ \{h_t\}\ )\ \ =\ \ \prod_{t=2}^{t_f} \varphi_{\sigma_h}(\ \tilde{H}_t + \tilde{C}_t + \tilde{V}_t\ -\ \tilde{h}_t\ ).$$

#### 3.5 Hospital Discharges

Data on hospital discharges are not age-structured in any of the data that we have. Therefore, the comparison between cumulative discharges in the data $u_t$ and cumulative discharges predicted by the ODE model $U_t(\theta)$ is modeled as

$$L_{10}\ \ =\ \ \prod_{t=2}^{t_f} \text{NegBin}(\ \tilde{u}_t\ ;\ \tilde{U}_t\ ,\ k_5\ )\ ,$$

with no "reporting rate" parameter or special accounting of missingness, other than time points being skipped in the calculation when two consecutive data points are not available.

#### 3.6 Likelihood of 11 passive surveillance data streams

The likelihood of all 11 passive surveillance data streams is the product of $L_1$ through $L_{10}$, and this likelihood $L$ is expressed given the ODE parameters ($\theta$), two reporting rates, five dispersion parameters for count data, and three error variances for occupancy data in the hospital, ICU, and on ventilation:

$$L(\text{ 11 data streams } \mid \theta, \rho, \rho_H, k_1, \ldots, k_5, \sigma_1, \sigma_3, \sigma_3\ )\ .$$

#### 3.7 Random Screening Data

In some instances, data are available that separate a department's asymptomatic/random screening data from their passive surveillance data. Passive surveillance is a health system confirming (by diagnostic test) self-reporting individuals who present with symptoms. Random screening is a health system planning testing campaigns in vulnerable populations, older populations, health-care workers, or certain high-contact groups. These individuals are tested randomly whether they have symptoms or not. Inference on the reporting parameter $\rho$ assumes that all data are collected via passive surveillance. If the random screening data are reported, the estimate of $\rho$ should be adjusted accordingly. Below we outline this adjustment for $\rho$ using Rhode Island's asymptomatic random screening data.

Let $c_t$ be the random screening coverage on day $t$, i.e. the fraction of the total population that receives a random test per day. For a symptomatic individual, we define $p_{\text{NCAS}}$ as the probability that this individual was not caught by random screening before symptoms occurred, at which point they would (with probability $\rho_t$) present to a hospital or clinic and be reported by the passive surveillance system. The key to clean integration of the passive surveillance and random screening testing components into a single likelihood is to avoid double-counting of individuals.

Table S2: COVID-19 tests administered in Rhode Island broken down by tested population. Data provided by Rhode Island Department of Health. Note that Public Symptomatic and Public Asymptomatic were estimated based on testing site location and not based on patient interviews.

| | Public Symptomatic | Congregate Care Resident | Congregate Care Employee | Public Asymptomatic |
|---|---|---|---|---|
| March | 3945 | 427 | 0 | 1 |
| April | 52,006 | 11,963 | 1106 | 279 |
| May | 57,124 | 24,265 | 9203 | 5816 |
| June | 49,195 | 23,803 | 12,737 | 6432 |
| July | 60,042 | 24,533 | 19,174 | 8372 |

Assuming a test sensitivity of $\sigma_E = 0.8$ and an incubation period duration of $\varepsilon = 6.0$ days, then

$$p_{\text{NCAS}} \;=\; (1 - \sigma_E c_t)^{\varepsilon} \tag{S16}$$

and the expected number of daily cases caught by passive surveillance is

$$\rho_t \cdot \tilde{J}_t \cdot p_{\text{NCAS}} \tag{S17}$$

which then becomes the mean of the negative binomial observation function of new cases, and the likelihood equation should now reflect that a negative binomial distribution with mean as above should be equal to $b_{p,t}$ = total number of positive cases reported through the passive surveillance system (but *not* the total number of cases reported for that day).

When $b_{p,t}$ is not available, we simply model the sum total of cases that we would expect to find through passive surveillance and random screening combined (i.e. the total number of new cases reported in a day). This expected value is

$$\rho_t \cdot \tilde{J}_t \cdot p_{\text{NCAS}} \quad + \quad N \cdot c_t \cdot \pi_t \tag{S18}$$

where $N \cdot c_t$ is the number of individuals tested in a day (through random screening) and $\pi_t$ is the probability that a randomly screened individual will test positive, which is simply the prevalence in the population:

$$\pi_t \;=\; \frac{\sigma_E E + \sigma_A A + \sigma_I I}{S + E + A + (1-\rho_t)I + (p_{\text{asymp}} + (1-\rho_t)p_{\text{symp}})R} \;. \tag{S19}$$

Above, we exclude all hospitalized cases as these individuals would not have an opportunity to take part in a random screening program, and we make sure to not double count individuals who had reported through passive surveillance and were informed of their positive test results.

Using the data in Table S2, we assume these age groups are: all ages, 70+, 20-70, and 20-70 (from left to right). From this we have that the *daily* number of random-screening tests (RST) by month and by age group was

$$\begin{aligned}
\text{RST}_{\text{March},20-70} &= 0.0 \\
\text{RST}_{\text{April},20-70} &= 46.2 \\
\text{RST}_{\text{May},20-70} &= 484.5 \\
\text{RST}_{\text{June},20-70} &= 639.0 \\
\text{RST}_{\text{July},20-70} &= 888.6
\end{aligned}$$

and the daily coverage ($c_t$) value for this entire age range is obtained by dividing the numbers above by 684,347 (population of the 20-70 age group in RI. For the 70+ age group this is, the daily RST numbers are

$$\begin{aligned}
\text{RST}_{\text{March},70+} &= 13.8 \\
\text{RST}_{\text{April},70+} &= 398.8 \\
\text{RST}_{\text{May},70+} &= 782.7 \\
\text{RST}_{\text{June},70+} &= 793.4 \\
\text{RST}_{\text{July},70+} &= 791.4
\end{aligned}$$

and we divide each of these numbers by 134,539 to get the daily coverage of random screening in the 70+ group

##### 3.8 Parameter Models

The ODE model described in section 2 takes a number of parameters as inputs. For almost all parameters we assign uniform prior distributions with values defined either by allowable limits or by reported estimates in the existing literature. Parameters that control hospitalization rates and length of hospital and ICU stays are given uniform priors that include values reported in the existing literature, but allow for variation in these rates across states.

The contact rate parameter $\beta_t$ controls the rate of contact between individuals, and is allowed to vary over time through a penalized spline expansion. We use a cubic B-spline expansion with one knot every seven days. If $s_\ell(t)$ is the $\ell$-th B-spline basis function evaluated at time $t$, then

$$\beta_t = \sum_{\ell=1}^{L} \alpha_\ell s_\ell(t) \tag{S20}$$

with $\alpha_1, \ldots, \alpha_L$ being the spline basis function loadings, which are parameters which need to be estimated. To model temporal consistency in contact rate parameters, we specify a correlated multivariate normal prior distribution on $\alpha_1, \ldots, \alpha_L$, with correlation defined to penalize second differences between the loadings $\alpha_1, \ldots, \alpha_L$. The variance of the multivariate normal prior is controlled by a variance parameter, $\sigma^2_\beta$, which is given an inverse gamma prior. Together, this hierarchical prior provides a flexible model for changing contact rates over time, with smoothness of the contact rates estimated from the data.

For the Rhode Island and Pennsylvania analyses, the reporting rate parameter $\rho_t$ is modeled as a penalized I-spline expansion, with knot locations chosen to allow for flexible, monotonic increases in the reporting rate from March to mid-June. The spline basis function loadings are estimated, with the same prior as described for the contact rate parameter $\beta_t$. However, in the Massachusetts analysis, the reporting rate $\rho_t$ is kept constant, with a uniform prior with limits between 0 and 1, as an I-spline results in a worse fit.

Parameters controlling the variance of data models around the mean defined by the ODE model come in two forms. Observed hospitalization data are given Gaussian likelihoods $(L_3, L_4, L_5)$ with variance parameters to be estimated by the data. These variance parameters are assigned diffuse inverse-gamma priors. Observed count data are given negative binomial likelihoods $(L_1, L_2)$ and have dispersion parameters which are assigned diffuse exponential priors.

##### 3.9 Inference

Inference on parameters controlling the ODE model, such as contact rate parameters and hospitalization stay parameters, as well as parameters that control variation in the observed data around ODE model means, is conducted under a Bayesian approach, with samples from the posterior distribution of model parameters obtained using a Markov chain Monte Carlo (MCMC) algorithm. All parameters that affect the underlying ODE model are proposed jointly. All other parameters are proposed one at a time. Missing values in age-structured new case counts and age-structured new hospitalization counts are assumed to be missing completely at random. Five separate Markov chains are run, and convergence is assessed visually.

#### 4 Parameters and Priors for Final Runs

The most challenging part of the inference was identification of the time from symptoms appearance to hospital admission (`time-symp-to-hosp`). This parameter was estimated in the early part of the epidemic in China as 4.64 days [4], 7 days [27], and 9.1 days [18]. Later estimates from the US and Europe put these estimates at between 3.5 to 7.0 days [2, 10, 11, 21]. Clinicians working with COVID-19 hospital admissions in Rhode Island indicated that the lower part of this range was more believable.

##### 4.1 Rhode Island

All 11 data streams from section 1.1 were included. Reporting rate $\rho$ was fit with an I-spline as RIDOH informed us that March reporting was very low due to lack of testing. Fit was substantially improved when comparing I-spline to constant reporting. Time from symptoms to hospitalization was given a prior of [2.0, 3.5] days as there was nothing in the data to inform this parameter and it appeared to have poor identifiability in many runs. Due to the completeness of the data in Rhode Island, there was strong support for age patterns changing (piece-wise constant assumption) from the spring phase to the summer phase, and there was strong support for a lower rate of ICU admission in the summer phase, although the effect size in RI for this was small. Posterior distributions for clinical and contact parameters are shown in Figure S5.

The prior distribution of the clinical parameters was constrained, with positive support limited to parameter combinations in which the expected cumulative reported symptomatic count total was within 10% of the observed cumulative total in the data. Thus, the MCMC sampler rejected parameter combinations in which the fitted cumulative case count deviated too far from the observed count.

#### 4.2 Massachusetts

Only 8 of 11 data streams from section 1.1 were included. Hospital deaths and hospital discharges were not available in Massachusetts, and the 'cumulative hospitalized' data stream could not be used because it was obtained by following up with symptomatic patients to ask about subsequent hospitalization (i.e. this was a fraction of the fraction $\rho$, and not a fraction of the total population $N$). Therefore, in order to obtain reliable fits and convergence in MCMC, probability of hospitalization by age was restricted to tight bounds around the median values obtained for the same parameters in the final RI run. In addition, the prior distribution on the length of the hospital stay (LOS) was set to be between 11.8 and 12.8 days, as this provided the best fit (lowest DIC) among several narrow prior ranges that were investigated. The LOS here refers to a typical medical-floor hospital stay for an individual that does not progress to critical care (these estimates are sometimes difficult to obtain as many studies mix ICU and non-ICU patients when reporting these averages). The prior distribution of the clinical parameters was constrained, with positive support limited to parameter combinations in which the expected cumulative reported symptomatic count total was within 10% of the observed cumulative total in the data. Thus, the MCMC sampler rejected parameter combinations in which the fitted cumulative case count deviated too far from the observed count.

Reporting rate $\rho$ was kept constant as an I-spline provided a worse fit. Time from symptoms to hospitalization was given a prior of [3.5, 5.0] days as, again, there was nothing in the data to inform this parameter. A mode of about 3.8 days for this parameter showed some identifiability in MA. The 'current number of patients on ventilator' data stream was consistently under-estimated in MA, and the parameters describing probability of progress from ICU to ventilation and duration on a ventilator kept being pulled to lower values by the inference, suggesting that there is something wrong in one of (1) model of clinical progression from hospital to ICU to ICU-on-ventilation, (2) likelihood function linking these data to the model, (3) the completeness of the 'currently on ventilator' data stream, or (4) the assumption that the pattern of clinical progression was constant throughout the epidemic.

In MA, the lockdown end-day had to be fixed at May 11 (plausible based on re-opening schedule of MA). Otherwise, the inference identified a second (lower likelihood) lockdown end-day in late May. Posterior distributions for clinical and contact parameters are shown in Figure S6.

#### 4.3 Pennsylvania

Only 9 of 11 data streams from section 1.1 were included. Hospital discharges were not available, and current number of patients in ICU was not available. Cumulative hospitalization data stopped being reported on the PA DOH website on May 21, and the inference for the reporting rate $\rho$ is largely influenced by these first 2.5 months of data. Reporting rate $\rho$ was fit with an I-spline (as in RI) as this provided a better fit than a constant reporting rate. Time from symptoms to hospitalization was given a narrow prior of [2.5, 3.0] days as there was nothing in the data to inform this parameter and it appeared to have poor identifiability in many runs. There was strong support for age patterns changing (piece-wise constant assumption) from the spring phase to the summer phase, and there was strong support for a much lower rate of ICU admission in the summer phase.

Hospital reporting appears to be incomplete in PA, i.e. the cumulative hospitalizations data stream reported through May 21 appeared to undercount new hospitalizations. Assuming that deaths are not undercounted, and that hospital occupancy and ventilator occupancy are not undercounted, a hospital reporting rate was fit and found to be identifiable. We estimate that 77.0% (95% CI: 62.8% – 87.1%) of new hospitalizations were being reported at this time. Posterior distributions for clinical and contact parameters are shown in Figure S7.

### References

[1] Safegraph documentation places manual. URL https://docs.safegraph.com/docs/places-manual.

[2] Matt Arentz, Eric Yim, Lindy Klaff, Sharukh Lokhandwala, Francis X. Riedo, Maria Chong, and Melissa Lee. Characteristics and Outcomes of 21 Critically Ill Patients With COVID-19 in Washington State. *JAMA*, 323 (16):1612–1614, April 2020. ISSN 0098-7484. doi: 10.1001/jama.2020.4326.

[3] Pavan K. Bhatraju, Bijan J. Ghassemieh, Michelle Nichols, Richard Kim, Keith R. Jerome, Arun K. Nalla, Alexander L. Greninger, Sudhakar Pipavath, Mark M. Wurfel, Laura Evans, et al. Covid-19 in Critically Ill Patients in the Seattle Region — Case Series. *New England Journal of Medicine*, 382(21):2012–2022, May 2020. ISSN 0028-4793, 1533-4406. doi: 10.1056/NEJMoa2004500.

[4] Qifang Bi, Yongsheng Wu, Shujiang Mei, Chenfei Ye, Xuan Zou, Zhen Zhang, Xiaojian Liu, Lan Wei, Shaun A. Truelove, Tong Zhang, et al. Epidemiology and transmission of COVID-19 in 391 cases and 1286 of their close contacts in Shenzhen, China: a retrospective cohort study. *The Lancet Infectious Diseases*, 20(8):911 – 919, 2020. ISSN 1473-3099. doi: https://doi.org/10.1016/S1473-3099(20)30287-5.

[5] Noelle Breslin, Caitlin Baptiste, Cynthia Gyamfi-Bannerman, Russell Miller, Rebecca Martinez, Kyra Bernstein, Laurence Ring, Ruth Landau, Stephanie Purisch, Alexander M. Friedman, et al. Coronavirus disease 2019 infection among asymptomatic and symptomatic pregnant women: two weeks of confirmed presentations to an affiliated pair of New York City hospitals. *American Journal of Obstetrics & Gynecology MFM*, 2(2):100118, May 2020. ISSN 25899333. doi: 10.1016/j.ajogmf.2020.100118.

[6] US Census Bureau. Glossary. URL https://www.census.gov/programs-surveys/geography/about/glossary.html. Section: Government.

[24] Henrik Salje, Cécile Tran Kiem, Noémie Lefrancq, Noémie Courtejoie, Paolo Bosetti, Juliette Paireau, Alessio Andronico, Nathanaël Hozé, Jehanne Richet, Claire-Lise Dubost, Yann Le Strat, Justin Lessler, Daniel Levy-Bruhl, Arnaud Fontanet, Lulla Opatowski, Pierre-Yves Boelle, and Simon Cauchemez. Estimating the burden of SARS-CoV-2 in France. page 5, 2020.

#### 5 Additional Figures

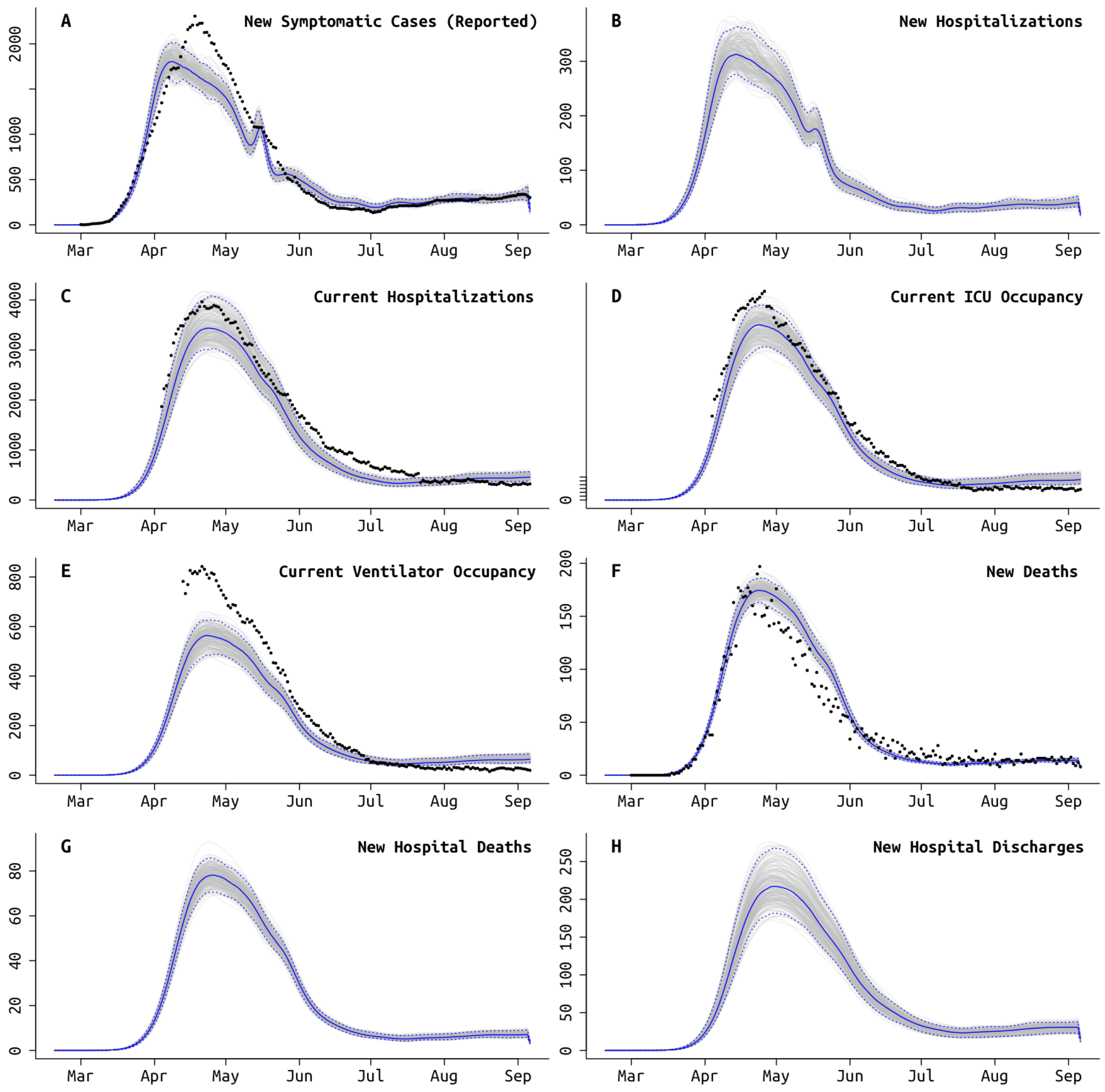

Figure S3: Model fit to Massachusetts daily data, using the best fit model which accounts for different age-based contact rates after the lockdown and a different rate of ICU admissions starting in early June (Table 3). Gray lines show 250 sampled trajectories from posterior, and blue lines are the median trajectories. Black circles are data points that show the daily (**A**) new reporting symptomatic cases, (**B**) new hospitalizations, (**C**) current number of patients hospitalized, (**D**) current number of patients in critical care, (**E**) current number of patients undergoing mechanical ventilation, (**F**) new deaths reported, (**G**) new hospital deaths reported, i.e. excluding deaths that occurred at home or at long-term care facilities, and (**H**) number of hospital discharges.

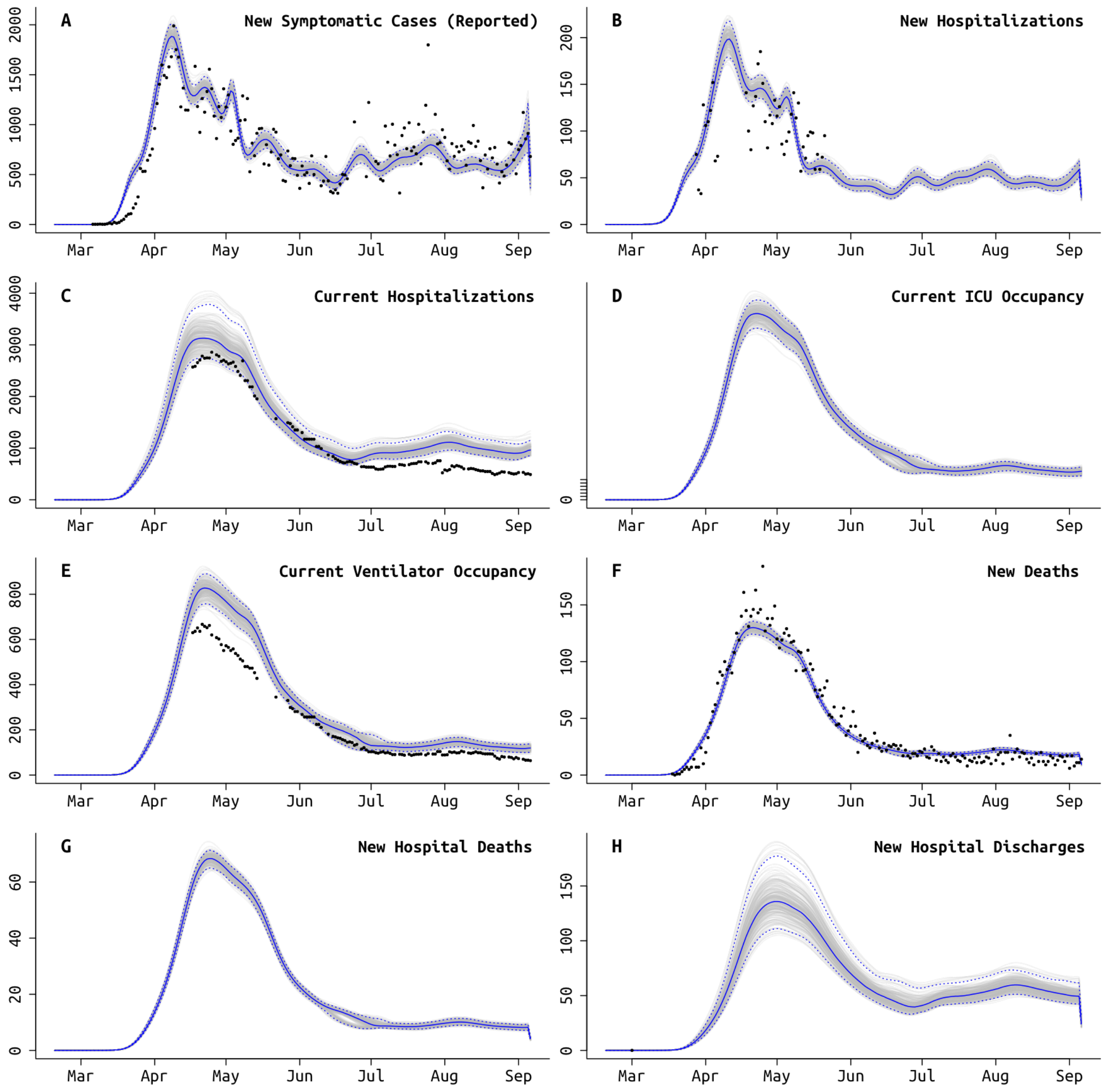

Figure S4: Model fit to Pennsylvania daily data, using the best fit model which accounts for different age-based contact rates after the lockdown and a different rate of ICU admissions starting in mid-June (Table 3). Gray lines show 250 sampled trajectories from posterior, and blue lines are the median trajectories. Black circles are data points that show the daily (**A**) new reporting symptomatic cases, (**B**) new hospitalizations, (**C**) current number of patients hospitalized, (**D**) current number of patients in critical care, (**E**) current number of patients undergoing mechanical ventilation, (**F**) new deaths reported, (**G**) new hospital deaths reported, i.e. excluding deaths that occurred at home or at long-term care facilities, and (**H**) number of hospital discharges.

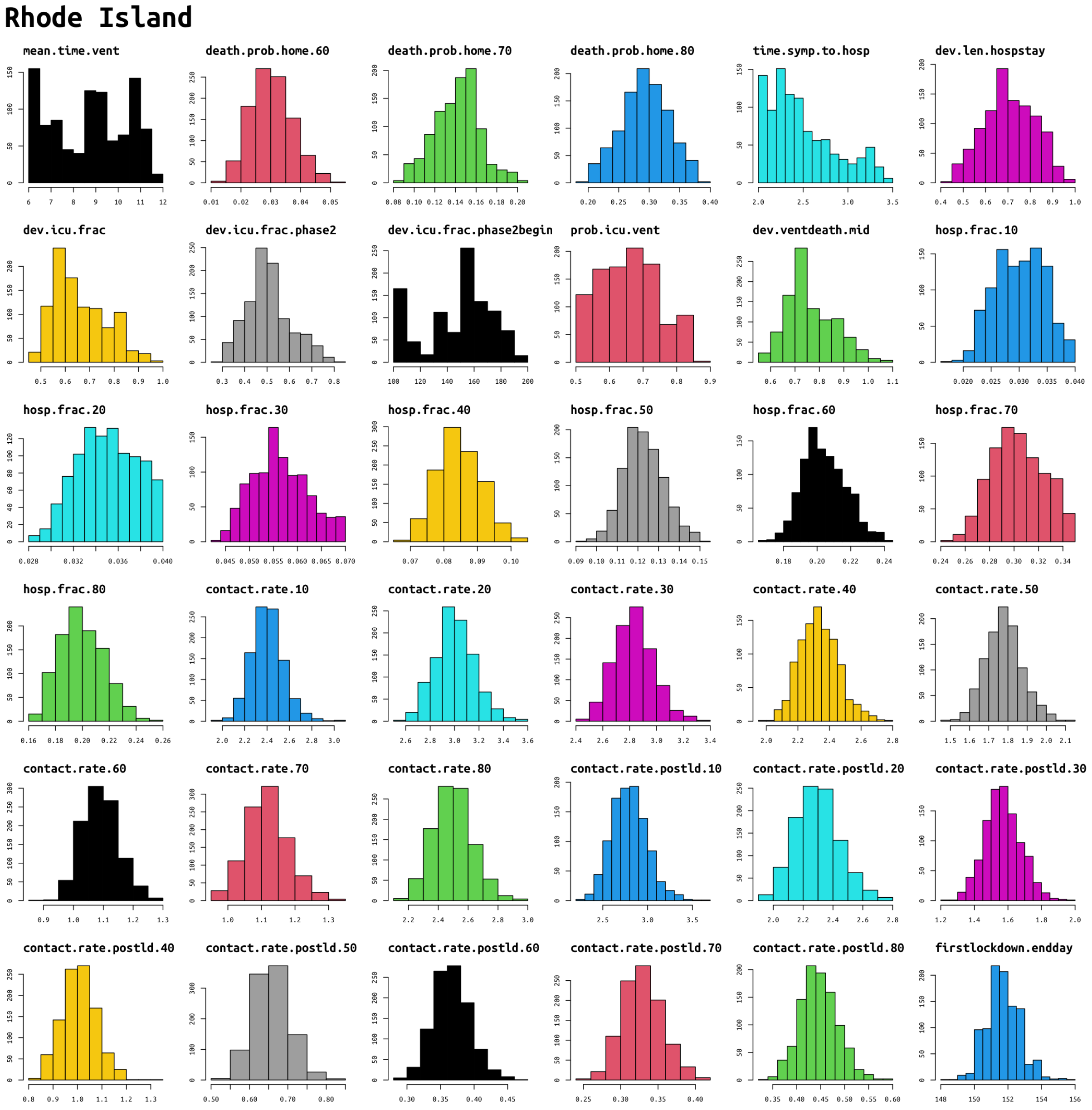

Figure S5: Posterior distributions for clinical and contact parameters found in the mathematical transmission model (see Section 2), inferred from the **Rhode Island** data. Histograms show 1000 posterior samples for each parameter, obtained via MCMC. The `dev.len.hospstay` scaling factor should be multiplied by 10.7 to get the length of a medical-floor hospital stay in days. The two `dev.icu.frac` parameters are scaling factors that are meant to be multiplied by the ICU admission probabilities in Lewnard et al [17]. The parameter `dev.ventdeath.mid` is the scaling factor $v$ in Table S1. Contact rates for age classes are presented as relative to the 0-9 age group. Hospitalization fraction (of symptomatics) is assumed to be the same for the 0-9 and 10-19 age groups.

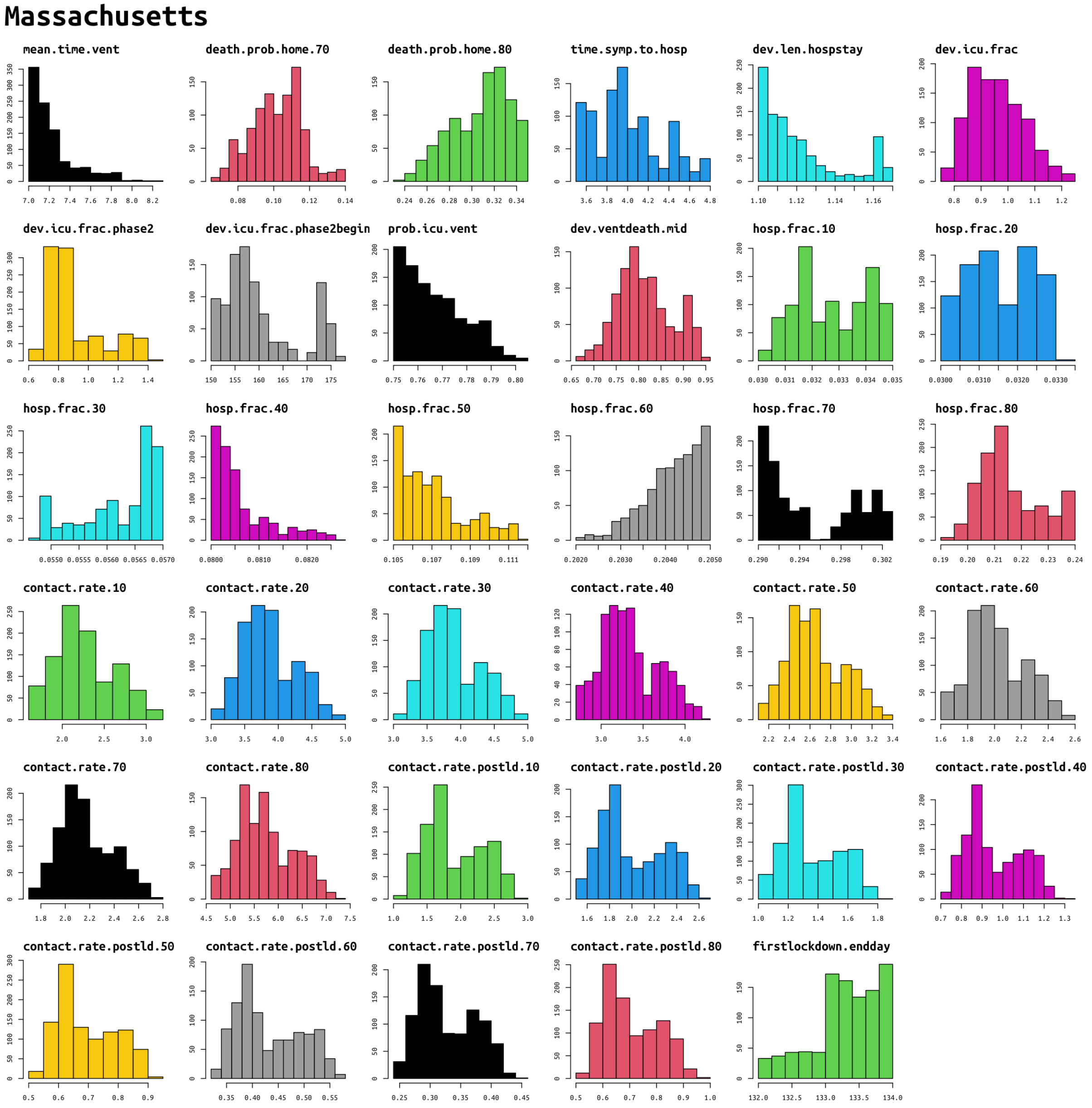

Figure S6: Posterior distributions for clinical and contact parameters found in the mathematical transmission model (see Section 2), inferred with the **Massachusetts** data. Histograms show 1000 posterior samples for each parameter, obtained via MCMC.

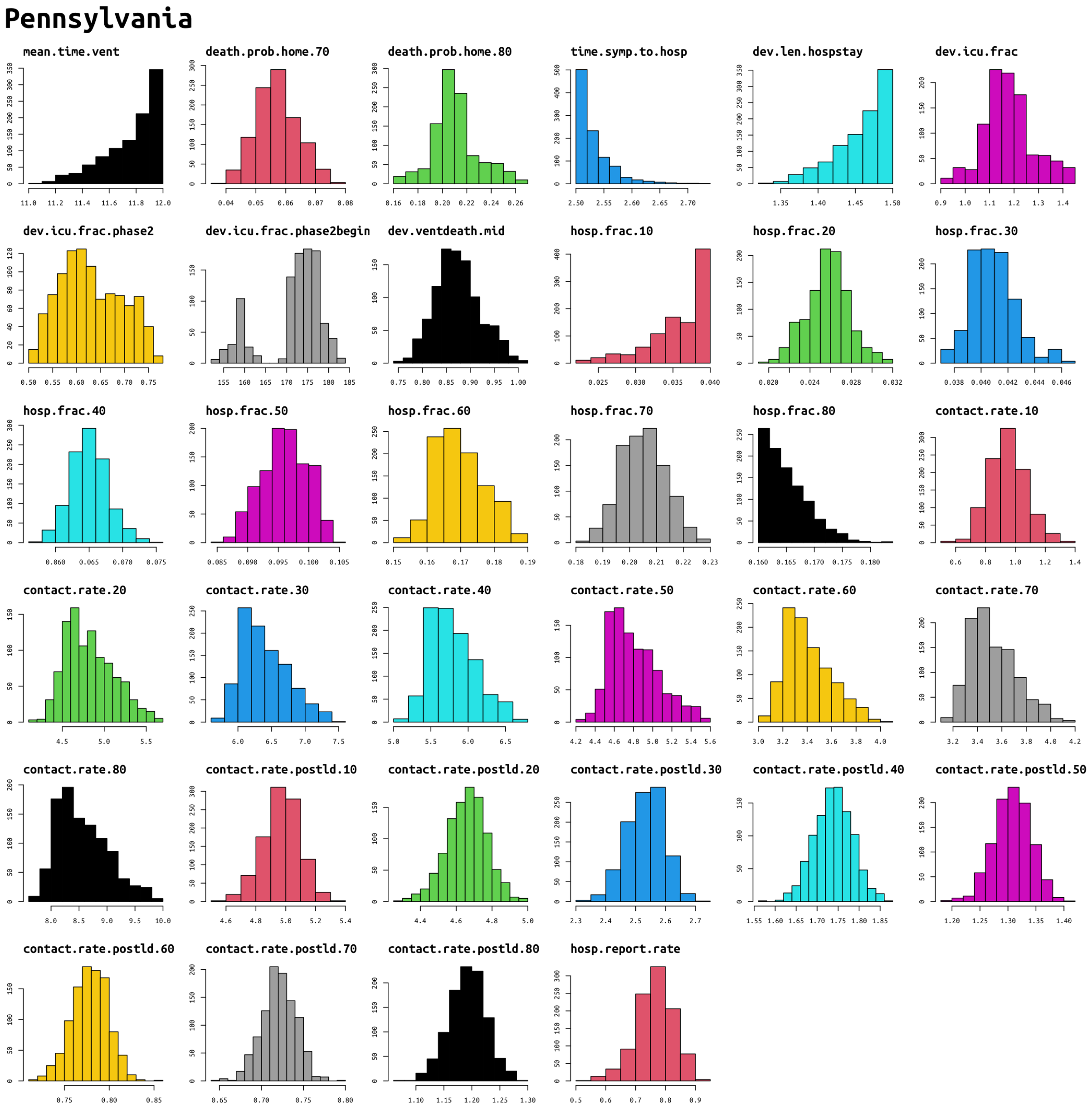

Figure S7: Posterior distributions for clinical and contact parameters found in the mathematical transmission model (see Section 2), inferred from the **Pennsylvania** data. Histograms show 1000 posterior samples for each parameter, obtained via MCMC. `hosp.report.rate` is the fraction of new hospitalizations that are reported.
